# Impact of lockdown on COVID-19 epidemic in Île-de-France and possible exit strategies

**DOI:** 10.1101/2020.04.13.20063933

**Authors:** Laura Di Domenico, Giulia Pullano, Chiara E. Sabbatini, Pierre-Yves Boëlle, Vittoria Colizza

## Abstract

**Background:** More than half of the global population is under strict forms of social distancing. Estimating the expected impact of lockdown and exit strategies is critical to inform decision makers on the management of the COVID-19 health crisis.

**Methods:** We use a stochastic age-structured transmission model integrating data on age profile and social contacts in Île-de-France to (i) assess the epidemic in the region, (ii) evaluate the impact of lockdown, and (iii) propose possible exit strategies and estimate their effectiveness. The model is calibrated to hospital admission data before lockdown. Interventions are modeled by reconstructing the associated changes in the contact matrices and informed by mobility reductions during lockdown evaluated from mobile phone data. Different types and durations of social distancing are simulated, including progressive and targeted strategies, with large-scale testing.

**Results:** We estimate the reproductive number at 3.18 [3.09, 3.24] (95% confidence interval) prior to lockdown and at 0.68 [0.66, 0.69] during lockdown, thanks to an 81% reduction of the average number of contacts. Model predictions capture the disease dynamics during lockdown, showing the epidemic curve reaching ICU system capacity, largely strengthened during the emergency, and slowly decreasing. Results suggest that physical contacts outside households were largely avoided during lockdown. Lifting the lockdown with no exit strategy would lead to a second wave overwhelming the healthcare system, if conditions return to normal. Extensive case-finding and isolation are required for social distancing strategies to gradually relax lockdown constraints.

**Conclusions:** As France experiences the first wave of COVID-19 pandemic in lockdown, intensive forms of social distancing are required in the upcoming months due to the currently low population immunity. Extensive case-finding and isolation would allow the partial release of the socio-economic pressure caused by extreme measures, while avoiding healthcare demand exceeding capacity. Response planning needs to urgently prioritize the logistics and capacity for these interventions.

## INTRODUCTION

More than half of the global population is under strict forms of social distancing^1,2^, with more than 90 countries in lockdown to fight against COVID-19 pandemic. France implemented the lockdown from March 17 to May 11, 2020^3^. The aim of this measure is to drastically increase the so-called social distance between individuals to break the chains of transmission and reduce COVID-19 spread. It is an unprecedented measure that was previously implemented only in Italy, Spain, Austria^2^, following the example of China^4^, to curb the dramatic increase of hospitalizations and admissions to ICU approaching saturation of the healthcare system.

The implementation of extreme measures of social distancing, including mobility restrictions, banning of mass gatherings, closure of schools and work activities, isolation and quarantine, helped control the first wave of COVID-19 pandemic in China^4–8^. Such exceptional coverage and intensive degree of intervention coupled with strict enforcement may be key to the resulting outcome. How this will play out in Europe is still uncertain^11,12^. Most importantly, how to relax such stringent constraints on social life and economy while controlling the health crisis remains under investigation^11–13^.

Here we use an age-structured mathematical model to (i) assess the current COVID-19 pandemic situation in France, (ii) evaluate the impact of the lockdown implemented nationwide on March 17, 2020, and (iii) estimate the effectiveness of possible exit strategies. The model is applied to the region of Île-de-France (heavily affected by the epidemic), it is data-driven and calibrated on hospital admission data for the region prior to lockdown. Different types and durations of social distancing interventions are explored, including a progressive lifting of the lockdown targeted on specific classes of individuals (e.g. allowing a larger portion of the population to go to work, while protecting the elderly), and large-scale testing for case-finding and isolation. The aim is to identify possible strategies to reduce the public health impact following the lifting of the lockdown.

The original version of this study was performed and made available as a preprint in mid-April, one month before the exit from lockdown. This revised version updates the comparison and validation of model projections, once data became available, while maintaining the context of the beginning of lockdown.

## METHODS

We consider a stochastic discrete age-structured epidemic model based on demographic and age profile data^14^ of the region of Île-de-France (**Figure 1**).

**Figure 1.**
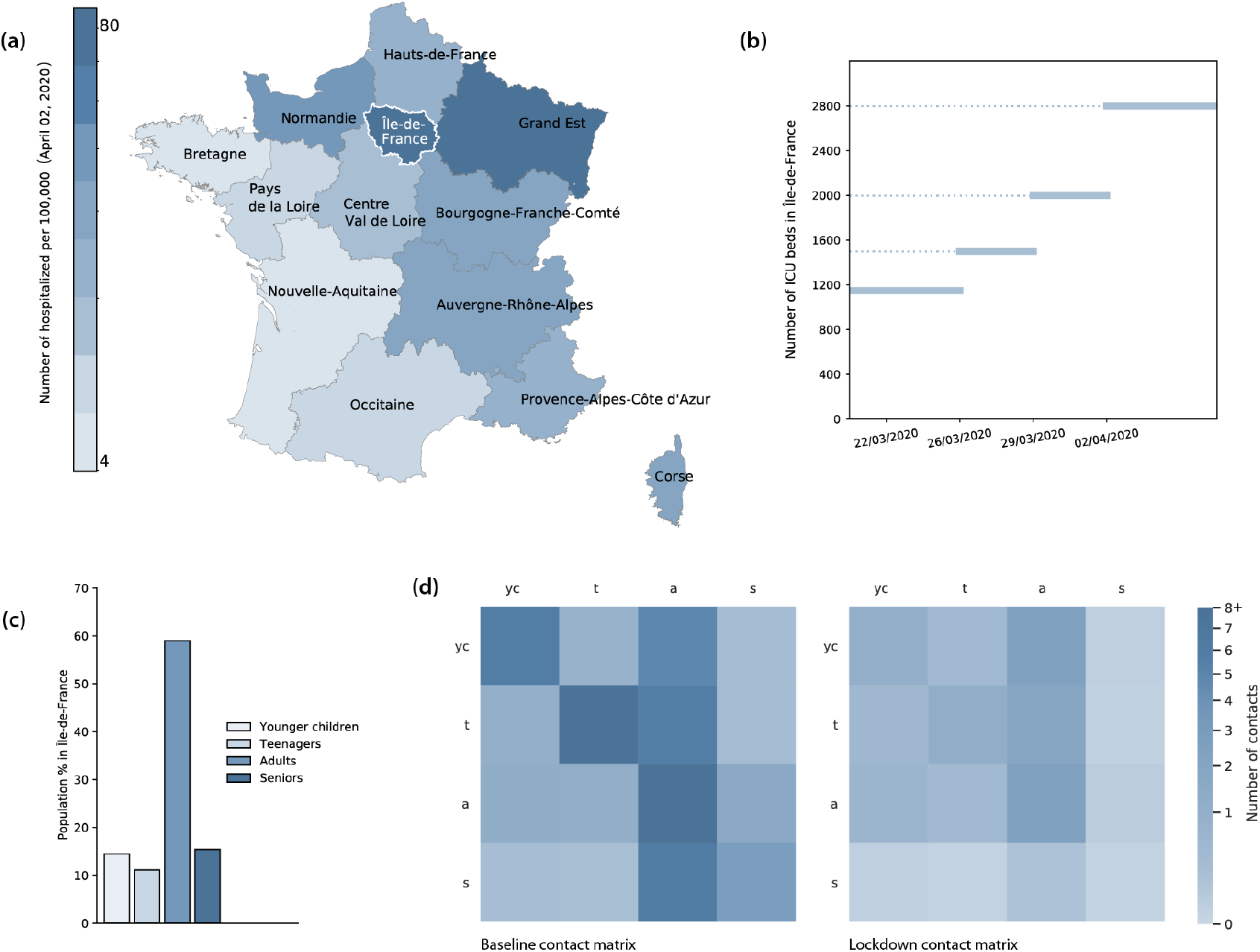
(a) Number of hospitalizations per 100,000 inhabitants per region in France as of April 2, 2020^15^. (b) Number of ICU beds in Île-de-France and increase of capacity over time^16^. (c) Age profile in Île-de-France region corresponding to younger children, teenagers, adults, seniors ([0,11), [11,19), [19,65), and 65+ years old, respectively). (b) Contact matrices in the baseline scenario (no intervention) obtained from data^17^ (left) and estimated for lockdown (right).

### Mixing

Four age classes are considered: [0,11), [11,19), [19,65), and 65+ years old, called in the following younger children (*yc*), adolescents or teenagers (*t*), adults (*a*), and seniors (*s*), respectively. We use social contact matrices measured in France in 2012 through a social contact survey^17^. The matrices represent the mixing in the baseline scenario (no interventions) between individuals in these four age groups (**Figure 1**), depending on the type of activity and place where the contacts occur (household, school, workplace, transport, leisure, other). Intervention measures are modeled through modifications of the contact matrices (see below).

### Compartmental model and transmission

Transmission dynamics follows a compartmental scheme specific for COVID-19 (**Figure 2**), where individuals are divided into susceptible, exposed, infectious, hospitalized, in ICU, recovered, and deceased. The infectious phase is divided into two steps: a prodromic phase (*I*_*p*_) occurring before the end of the incubation period, followed by a phase where individuals may remain either asymptomatic (*I*_*a*_) or develop symptoms. In the latter case, we distinguish between different degrees of severity of symptoms, ranging from paucisymptomatic (*I*_*ps*_), to infectious individuals with mild (*I*_*ms*_) or severe (*I*_*ss*_) symptoms, according to data from Italian COVID-19 epidemic^18^ and estimates from individual-case data from China and other countries^19^. We explore two values of the probability of being asymptomatic, namely *p*_*a*_ =20% and 50%, in line with available estimates^20–22^. Individuals in the prodromic phase, asymptomatic and paucisymptomatic individuals have a smaller transmission rate with respect to individuals with moderate or severe symptoms, as reported by contact tracing investigations^23^ and estimated in Ref.^8^. Current evidence from household studies, contact tracing investigations, and modeling works suggest that children are as likely to be infected by COVID-19 as adults, but more likely to become either asymptomatic or paucisymptomatic^22,24–26^. This may explain the very small percentage (<5%) of children in COVID-19 confirmed cases worldwide^27^. Here we assume that children in both classes (younger children and adolescents) are equally susceptible as adults, following Ref.^24^, and that they become either asymptomatic or paucisymptomatic only. Different relative susceptibility or infectivity of children compared to adults are tested for sensitivity analysis.

**Figure 2.**
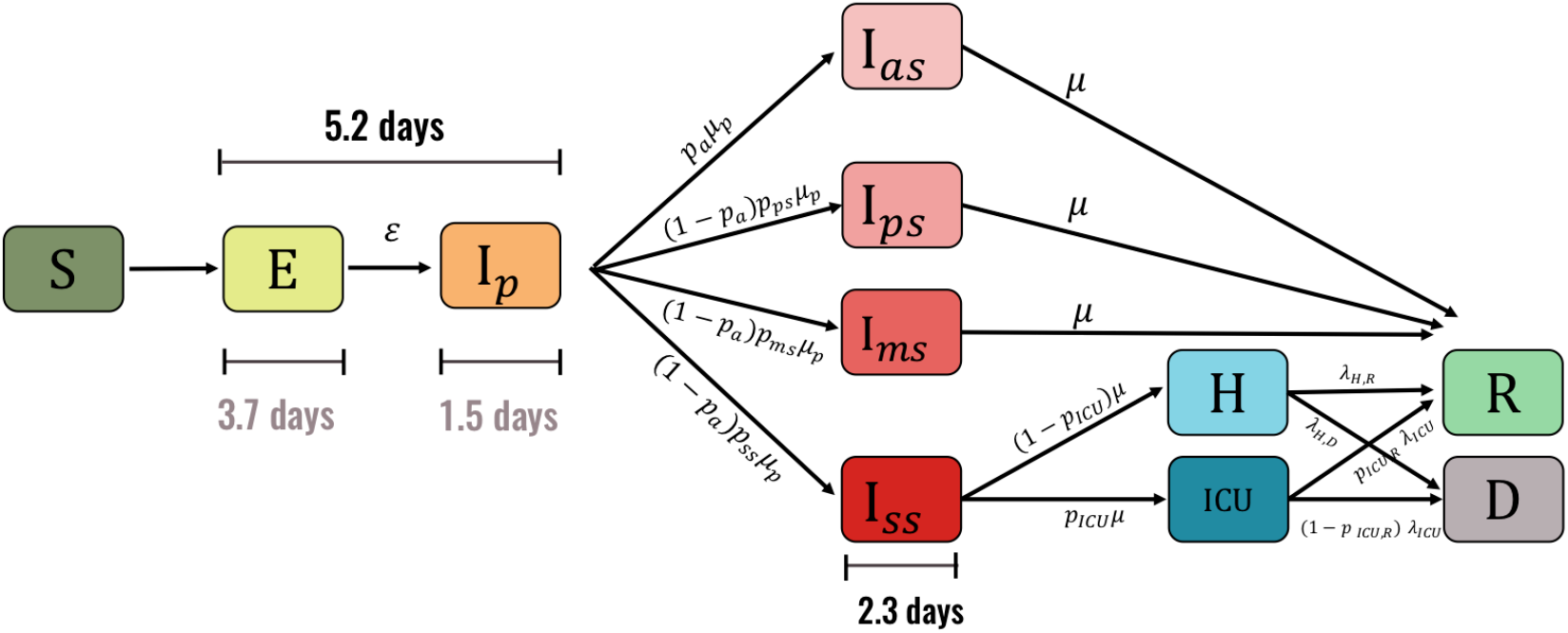
Compartmental model. S=Susceptible, E=Exposed, I_p_= Infectious in the prodromic phase (the length of time including E and I_p_ stages is the incubation period), I_a_=Asymptomatic Infectious, I_ps_=Paucysymptomatic Infectious, I_ms_=Symptomatic Infectious with mild symptoms, I_ss_=Symptomatic Infectious with severe symptoms, ICU=severe case admitted to ICU, H=severe case admitted to the hospital but not in intensive care, R=Recovered, D=Deceased.

The compartmental model includes hospitalization and admission to ICU for severe cases. ICU admission rates, hospital case fatality, lengths of stay after admission are informed from French hospital data for patient trajectories in Île-de-France region (SIVIC database maintained by the Agence du Numérique en Santé and Santé Publique France^28,29^) (see also Additional file 1). ICU beds occupation is the indicator used to evaluate the capacity of the region to face the surge of patients requiring intensive care. Since we do not use hospital beds occupation for this evaluation, we neglect the time spent in the hospital after exiting intensive care.

Parameters, values, and sources used to define the compartmental model are listed in Table S1 of the Additional file 1^8,18,19,21,28,30–35^.

### Change of behavior due to severe illness

We assume that infectious individuals with severe symptoms reduce of 75% their number of contacts because of the illness they experience, as observed during 2009 H1N1 pandemic^36^. Higher reductions are tested as possible interventions of self-isolation (see below).

### Social distancing interventions

We implement social distancing interventions by reconstructing the associated changes of the contact matrices, accounting for a reduction of the number of contacts engaged in specific settings. More precisely:

- *School closure:* the contact matrix for school is removed. We consider that 5% of adults may stay at home to take care of children while schools are closed (not applied with telework or lockdown).
- *Telework performed by a given % of individuals:* contacts at work and on transports are reduced to account for the % of workers not going to work anymore. In France telework is performed daily by 6% of the active adult population^37^. Here we consider three values: 25%, 50% (as declared by participants of a crowdsourced system monitoring COVID-19 associated behaviors in France^38^), and 70%. The 70% reduction is considered during lockdown, informed by the reduction of mobility documented during the first weeks of lockdown, and resulting from the analysis of mobile phone data in the region^39^. The percentages associated to telework more generally include all individuals not going to their place of work (because they work remotely, they stopped working due to restrictions, or other conditions). Household contacts are increased proportionally to each adult staying at home based on statistics comparing weekend vs. weekday contacts^17^ and proportion of adults working during the weekend^40^.
- *Senior isolation:* contacts established by seniors are reduced by a given % to model a marked social distancing targeting only the age class at higher risk of complications. We consider 75% and 90%.
- *Banning of social events and closure of all non-essential activities:* Contacts established during leisure and other activities are completely or partially (50%) removed.
- *Case isolation:* in a scenario with large-scale and rapid testing availability, we consider that 25%, 50% or 75% of all infectious individuals (also including asymptomatic) reduce promptly their contacts by 90% throughout their illness as they self-isolate. This simulates the outcome of aggressive contact-tracing and isolation^31,41^. To account for a delay in tracing, testing and self-isolation, we consider that infected individuals in their prodromic stage maintain their contacts as in the no intervention scenario.

Combinations of the above social distancing interventions simulate the lockdown and alternative less strict options as exit strategies (**Table 1**). Physical contacts during lockdown are removed to account for the adoption of physical distancing; an alternative version of the matrix maintaining physical contacts during lockdown is tested for sensitivity. We explore possible progressive exit of specific categories of individuals from lockdown (e.g. gradually reopening some businesses while still requiring a high rate of telework where possible), while maintaining strict social distancing for those at higher risk of complications (e.g. protecting seniors through isolation), as well as increasing testing capacities over time. Results without physical contacts in the exit strategies are tested for sensitivity analysis.

**Table 1.** Combinations of social distancing interventions. Telework under intensive measures refers to the ensemble of individuals who remain at home (including teleworkers, but also those who do not work anymore because of the lockdown, to care for children, or other conditions).

|  |  | **School closure** | **Telework**<br>(individuals not going to work) | **Senior isolation** | **Closure non-essential activities** | **Case isolation** |
| --- | --- | --- | --- | --- | --- | --- |
| *Lockdown* |  | Yes; 100% contacts of children on transports removed | 70% <sup>39</sup> | Yes, with 90% contact reduction | Yes, 100% closure | No |
| *Set of strict interventions* |  | Yes; 100% contacts of children on transports removed | 50% <sup>38</sup> | Yes, with 75% contact reduction | Yes, 100% closure | No |
| *Set of moderate interventions* |  | Yes; 50% contacts of children on transports removed | 50% <sup>38</sup> | Yes, with 75% contact reduction | Yes, 50% closure | No |
| *Set of mild interventions* |  | Yes; contacts of children on transports are not removed | 25% | Yes, with 75% contact reduction | No | No |
| *School closure and senior isolation* |  | Yes; contacts of children on transports are not removed | As in baseline | Yes, with 75% contact reduction | No | No |
| *Lockdown + case isolation* |  | Yes; 100% contacts of children on transports removed | 70% <sup>39</sup> | Yes, with 90% contact reduction | Yes, 100% closure | Yes, for 50%, 75% of cases |
| *Set of strict interventions + case isolation* |  | Yes; 100% contacts of children on transports removed | 50% <sup>38</sup> | Yes, with 75% contact reduction | Yes, 100% closure | Yes, for 25%, 50%, 75% of cases |
| *Set of moderate interventions + case isolation* |  | Yes; 50% contacts of children on transports removed | 50% <sup>38</sup> | Yes, with 75% contact reduction | Yes, 50% closure | Yes, for 50% of cases |
| *Set of mild interventions + case isolation* |  | Yes; contacts of children on transports are not removed | 25% | Yes, with 75% contact reduction | No | Yes, for 50%, 75% of cases |

Intervention scenarios are based on the nationwide implementation of the lockdown in France from March 17, 2020 up to May 11. Timelines of explored scenarios are illustrated in **Figure 3**. Different durations of the lockdown (till the end of April, end of May) are also considered for sensitivity.

**Figure 3.**
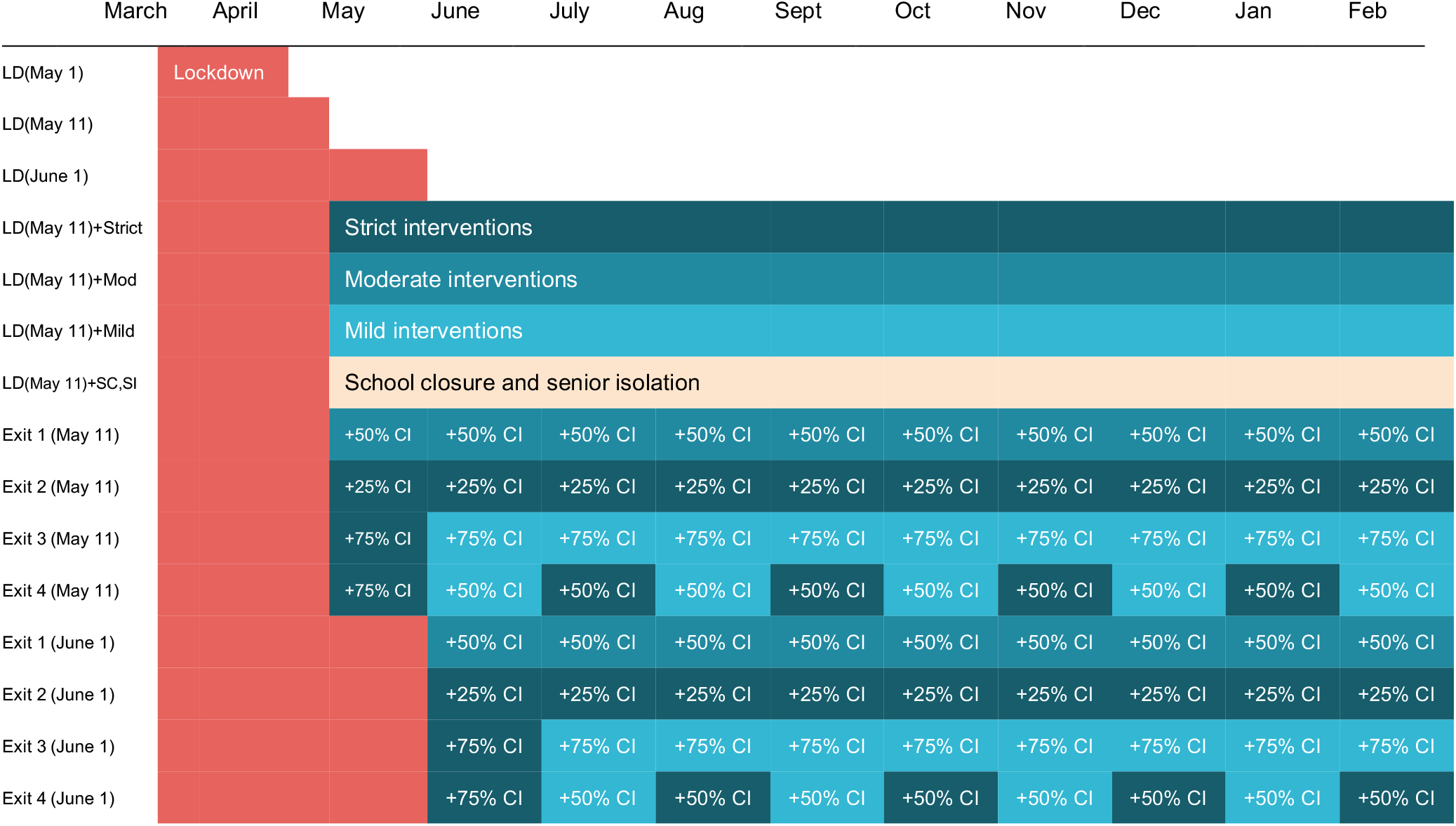
Scenarios (color code as in Table 1; CI refers to case isolation).

### Calibration and validation

The model is calibrated on hospital data^28^ specifying the number of COVID-19 positive hospital admissions in Île-de-France prior to lockdown. We use a maximum likelihood approach to fit the transmission rate per contact and the starting date of the simulation considering data in the interval March 1-23, 2020 (Additional file 1). Hospital admissions in the interval March 17-23 were included as still not affected by lockdown, due to the delay between date of infection and date of hospitalization (∼1 week). Parameters estimating the hospital trajectories of COVID-19 patients are based on data up to April 5.

We also retrospectively fit the epidemic trajectory during the lockdown phase, once data became available, to estimate the deviation from our assumptions (Additional file 1).

The simulated incidence of clinical cases (mild and severe symptoms) is compared to the regional incidence of COVID-19 cases estimated by the syndromic and virological surveillance system^29,42^ (see also Additional file 1) from week 12 (March 16 to 22, 2020). Validation is performed *a posteriori* on the epidemic trajectories of hospital admissions, ICU admissions, and ICU beds occupation, during the full lockdown period, once data became available.

### Evaluation

Each intervention of social distancing is compared to the no intervention scenario in terms of final attack rate, peak time, peak incidence, ICU beds demand in the region. The total number of ICU beds in Île-de-France has progressively increased over time up to a capacity of 2,800 ICU beds^43^ in an effort to sustain the first wave and respond to the emergency (**Figure 1**). Passed the first wave, we consider an ICU capacity restored at 1,500 beds in the months following the emergency^44^, as the emergency response is not sustainable in the long term. This capacity corresponds to a 25% increase compared to standard pre-COVID-19 size (1,147 beds), assuming that a certain level of preparedness is kept in the upcoming months to face the possible evolution of the pandemic.

For each scenario, we perform 250 stochastic runs, median curves are displayed together with the associated 95% probability ranges.

### Sensitivity analysis

Results reported in the main paper refer to *p*_*a*_ =0.2, those for *p*_*a*_ =0.5 are shown in the Additional file 1. We compare the lockdown based on our reconstructed matrix with (i) a less stringent lockdown under the reduction of contacts measured in the UK^45^, (ii) a more stringent lockdown under the reduction of contacts measured in China^46^, (iii) a fit of the model during the lockdown phase, (iv) an additional version of our reconstructed matrix where physical contacts are kept. Exit strategies *Exit 1-4* are also examined with a lockdown lifted at the beginning of May, or of June, and assuming that physical contacts are avoided in the upcoming months. In addition, we test different relative susceptibility and infectivity of children compared to adults. We did not consider scenarios assuming that infectiousness is constant across asymptomatic and symptomatic individuals, as in contrast with available evidence^8,23^.

## RESULTS

### Reproduction number, start of the epidemic, population infected

The reproductive number for our model is estimated to be *R*_*a*_=3.18 [3.09, 3.24] (95% confidence interval), computed with the next-generation approach^47^ based on the transmission rate estimated from hospital admissions in Île-de-France prior to lockdown (**Figure 4**).

**Figure 4.**
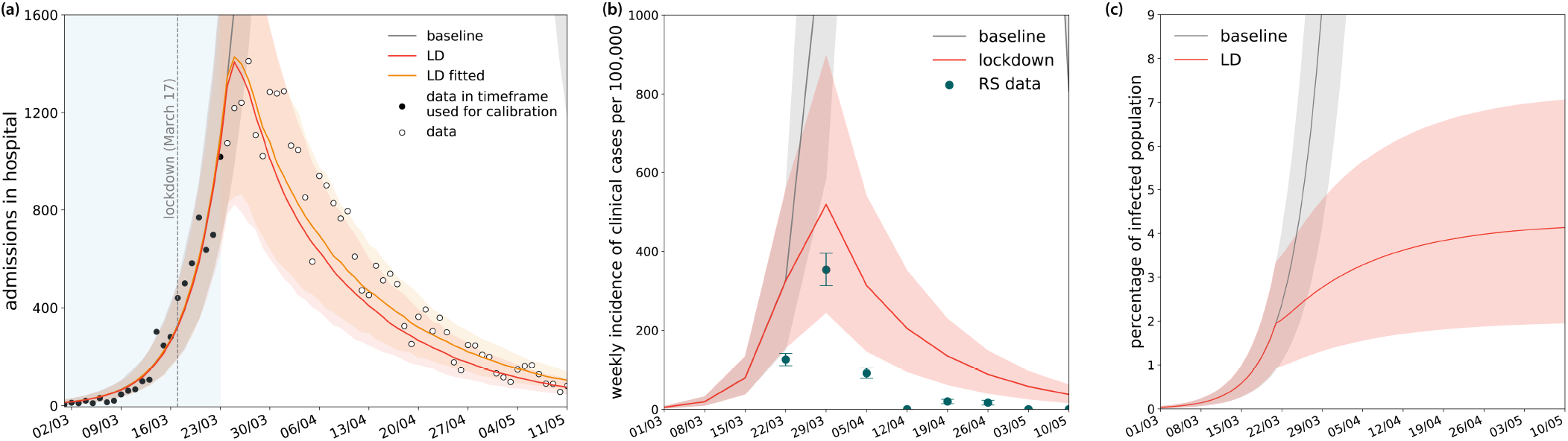
Calibration of the model and estimates of weekly incidence and percentage of population infected. (a) Calibration of the model on data of daily hospital admissions in Île-de-France prior to lockdown, and projections for the lockdown phase. Black dots indicate data in the timeframe used for calibration, also indicated by the region in light blue; white dots indicate data in the prediction timeframe. Our model predictions are compared to results obtained by fitting out model also in the lockdown phase (orange line for the median curve). (b) Simulated weekly incidence of clinical cases (mild and severe) compared to estimates of COVID-19 positive cases in the region provided by syndromic and virological surveillance (Reseau Sentinelles (RS) data)^42^. (c) Simulated percentage of population infected over time. Results are shown for *p*_*p*_=0.2. Shaded areas correspond to 95% CI.

Reported hospitalizations are consistent with an epidemic seeded in the region at the end of January 2020. The percentage of population in Île-de-France predicted to be infected on May 11 ranges from 2% to 13% considering both values of the probability of being asymptomatic (**Figure 4**, and Additional file 1 for the higher asymptomatic rate scenario). Overall infection fatality ratio is estimated to range from 0.7% to 1.2%.

### Lockdown followed by combination of interventions of different degrees of intensity

The changes in contact matrices reconstructed to simulate the social distancing measures implemented during lockdown reduce the number of contacts by 81% compared to baseline mixing patterns (**Figure 1**). This allows a substantial reduction of the reproductive number well below 1 (*R*_*LD*_=0.68 [0.66, 0.69]). Under these conditions, the predicted incidence of clinical cases slows down and reduces during lockdown, following the tendency reported by syndromic and virological surveillance (**Figure 4**). The overestimation of clinical cases is explained with a consultation rate estimated to be around 35% in the reported period, as documented from crowdsourced data^38^ (Fig. S3 in Appendix file 1). Fitting the model to the epidemic trajectory during the lockdown phase would require a correction of 5% increase in the transmissibility per contact (**Figure 4** and Additional file 1).

The predicted number of ICU beds occupied saturates towards the strengthened capacity in the region, before slowly decelerating and reducing over time. Predictions are consistent with observations (**Figure 5**). The median ICU demand slightly overestimates the data, as our model does not account for the transfer of patients in intensive care to less affected regions.

**Figure 5.**
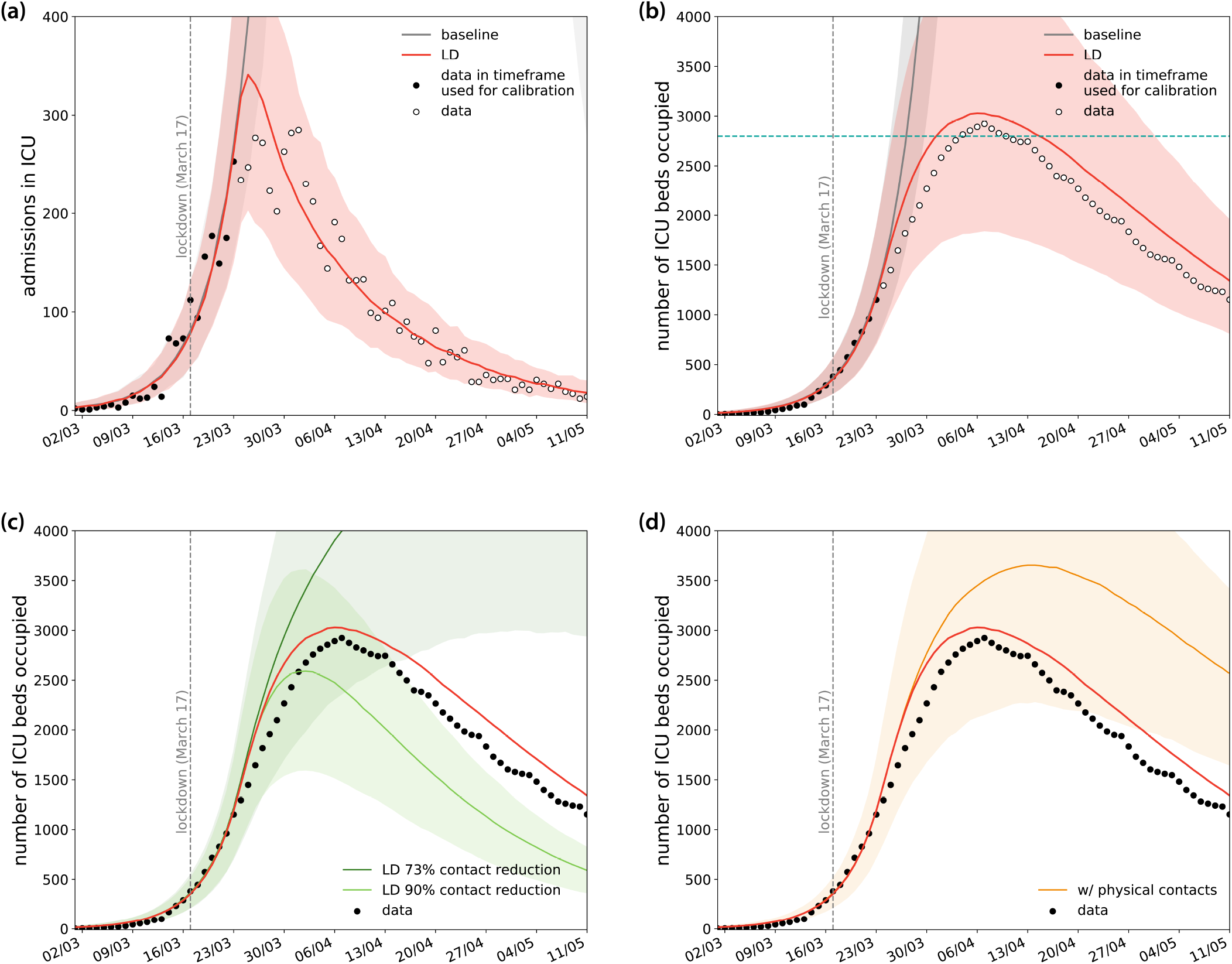
Lockdown projections compared to data. (a) Simulated daily incidence of admissions in ICU over time. (b) Simulated number of ICU beds occupied during lockdown. In panels (a) and (b) black dots indicate data in the timeframe used for calibration (fit to hospital admission data before lockdown, see Fig. 4) and white dots indicate data in the prediction timeframe. (c) Simulated number of ICU beds occupied assuming a less stringent lockdown, under the reduction of contacts measured in the UK^45^ (73%), and a more stringent lockdown, under the reduction of contacts measured in China^46^ (90%). The median prediction of our model is also shown for comparison (red curve). (d) Simulated number of ICU beds occupied resulting from considering the inclusion of physical contacts during lockdown. The median prediction of our model is also shown for comparison (red curve). In all plots: vertical dashed line refers to the start of the lockdown; horizontal lines refer to ICU capacity in the region (see Fig1b); shaded areas correspond to 95% probability ranges. Results are shown for *p*_*p*_=0.2.

Model projections indicate that by May 11 the region would experience 516 [363, 682] new clinical cases per day (corresponding to 1,170 [808, 1534] new infections) and 75 [49, 107] new admissions in hospitals as shown in **Figure 4** (of which 18 [10, 30] in ICUs), with an ICU system occupied at 48% [31, 68]% of currently strengthened capacity (**Figure 5)**.

Assuming a 90% decrease during lockdown predicts faster decrease in bed occupation (**Figure 5**). With a less stringent reduction (73%), *R*_*LD*_ is just below 1 and the trend of occupied beds in ICUs is predicted to continue to increase up to end of April. None of these situations is consistent with the data. Considering physical contacts during lockdown would lead to a higher occupancy of ICU beds than observed.

Lifting the lockdown with no exit strategy, and going back to normal conditions, leads to a delay of the peak compared to the no-intervention scenario (delay of about the duration of the lockdown) with minimal effect on peak incidence (**Figure 6**). The peak number of ICU beds required is estimated to be more than 60 times the restored regional capacity if no strategy is implemented after lockdown.

**Figure 6.**
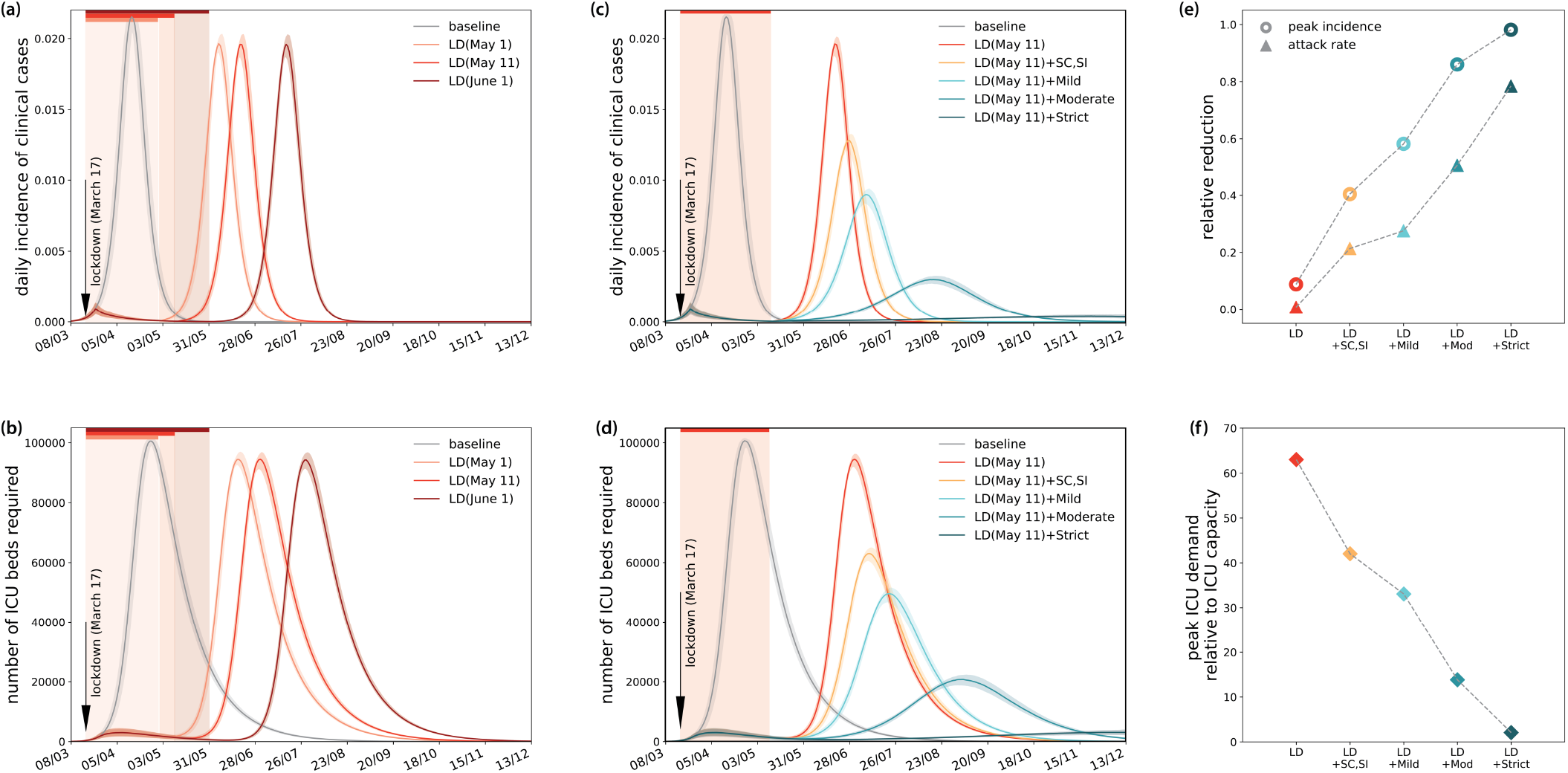
Simulated impact of lockdown of different durations and exit strategies. (a) Simulated daily incidence of clinical cases assuming lockdown till end of April, May 11, end of May. (b) Corresponding demand of ICU beds. (c) Simulated daily incidence of clinical cases assuming lockdown till May 11, followed by interventions of varying degree of intensity. (d) Corresponding demand of ICU beds. (e) Relative reduction of peak incidence and epidemic size after 1 year for each scenario. (f) Peak ICU demand relative to restored ICU capacity of the region (1,500 beds). In all panels, the color code is as in Table 1, and scenarios are identified as reported in Figure 3. Vertical colored areas indicate the time period of lockdown under the different measures. Baseline scenario corresponds to no intervention. Results are shown for *p*_*p*_=0.2. Shaded areas correspond to 95% probability ranges.

Combined interventions of different degrees of intensity implemented indefinitely after lifting the lockdown in May substantially delay and mitigate the epidemic (**Figure 6**). School closure coupled with senior isolation, and mild interventions reduce the peak incidence by approximately half (40%, 58% respectively). Interventions of moderate intensity or higher (i.e. schools are closed, 50% of active individuals work remotely, and at least 50% of non-essential activities are closed; seniors remain in isolation) suppress the peak with more than 80% reduction gaining significant delay compared to the no-exit strategy. Despite the strong mitigation of these scenarios, the peak demand on the healthcare system is predicted however to exceed capacity by a large amount if interventions are at most of moderate intensity (15 to 40 times higher than capacity) (**Figure 6**). Strict intervention would still require twice the restored capacity of the system during the second wave.

### Social distancing measures with case isolation

Implementing aggressive case-finding and isolation together with social distancing allow the release of the lockdown in May, engaging the ICU services below their maximum capacity throughout the epidemic (**Figure 7**). On the medium- to long-term, different intensity of social distancing interventions can be maintained, depending on the testing and isolation capacity. For example, strict interventions if only 25% of cases are promptly identified and isolated (*Exit 2*), or moderate interventions if such capacity is increased (50% case isolation, *Exit 1*). These two scenarios predict the occupation of more than 500 beds in ICU up to the month of June. Lifting the lockdown at the beginning of May would maintain ICU occupation over this level for the entire summer (Additional file 1). Delaying till early June would achieve a stronger suppression of the epidemic.

**Figure 7.**
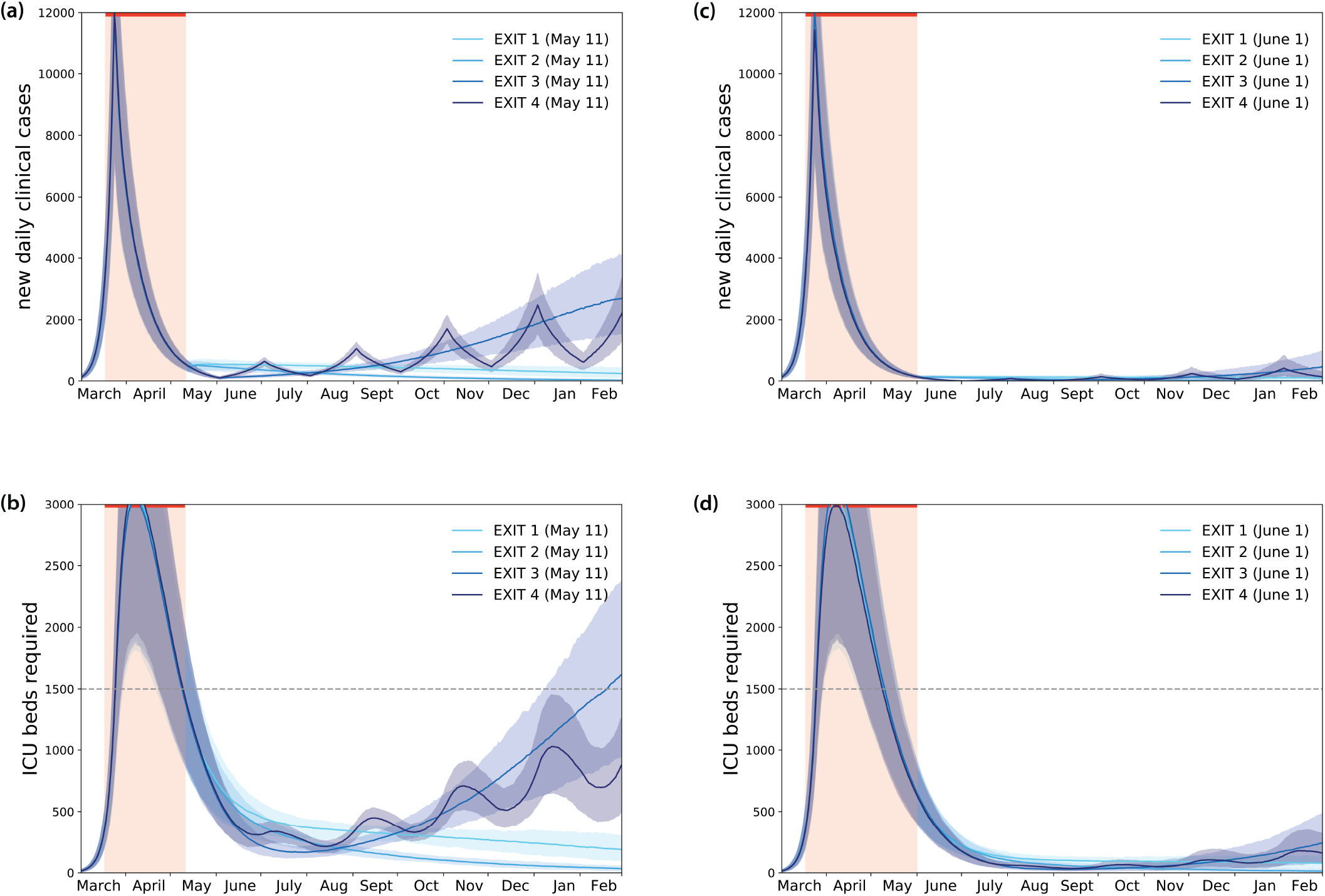
Simulated impact of lockdown and exit strategies with large-scale testing and case isolation. (a) Simulated daily new number of clinical cases assuming the progressive exit strategies illustrated in Figure 3. (b) Corresponding demand of ICU beds. (c) As in (a) with exit strategies implemented after a lockdown till the end of May. (d) Corresponding demand of ICU beds.

Further relaxing social distancing constraints (e.g. allowing a larger proportion of individuals to go to work and the full reopening of activities) would be possible by progressively going through decreasing levels of intensity of interventions (from lockdown to strict interventions to mild interventions afterwards) while maintaining highly efficient tracing (*Exit 3*). Alternatively, strict and mild interventions can be rotated every month if case isolation capacity is less efficient (*Exit 4*).

All these scenarios foresee, however, that schools are closed, and seniors remain isolated.

All exit strategies would be able to maintain the epidemic under control in the upcoming months if a larger proportion of infected individuals is asymptomatic. This is due to the smaller fraction of individuals in the population with severe symptoms requiring hospitalization (Additional file 1). If younger children and adolescents are less susceptible than older age classes, transmissions would mainly occur from infectious adults and seniors. A higher transmission rate, with respect to the equal susceptibility scenario, is thus required to reproduce the epidemic trajectory prior to lockdown. This is due to the fact that younger age classes have a large number of contacts, but these contacts become less important in the disease spread if individuals are less susceptible to contract the infection. In the scenario analyses, the reduced susceptibility assumption would lead on average to a higher flux of hospital admissions, as older age classes are the ones with higher probability of being hospitalized.

Exit strategies however exist that would maintain the epidemic under control (Additional file 1). Also, changes in the risk of transmission from younger children would lead to similar results. If individuals continue to avoid all physical contacts in the next months, all exit strategies considered here foresee the suppression of the epidemic in the region (Additional file 1).

## DISCUSSION

We use a stochastic age-structured epidemic transmission model calibrated on hospital admission data in Île-de-France to evaluate the impact of lockdown and exit strategies in controlling COVID-19 epidemic in the region. Our estimate of the reproductive number prior to lockdown is in line with estimates for the epidemic growth in Europe prior to the implementation of interventions^9-10^, with results from a meta-analysis of the literature^12,45^, and with concurrent studies in France^35,48^. We predict it decreased significantly during lockdown with a 95% probability range well below 1. Under these conditions, the occupancy of the ICU system reaches a plateau before clearly showing a decreasing tendency after several weeks of lockdown, as also observed in other countries^18^. As of May 11, the model predicts a 48% of ICU occupancy, with less than 30 new admission to ICUs per day.

Lifting lockdown with no exit strategy in place, i.e. going back to pre-lockdown conditions, would inevitably lead to large rebound effects, as the immunity of the population is estimated to be still very low (from 2% to 13% considering both values of probability of being asymptomatic explored), in agreement with other estimates^9,35^. Prolonged interventions of moderate to high intensity could additionally delay the epidemic peak by at least 2 months compared to the no-exit strategy and reduce its peak incidence by more than 80%, but would not avoid exceeding ICU capacity (peak demand of 2-15 times the restored ICU capacity of the region). Even with a 100% increase of ICU capacity to face the second peak, strict interventions would be required for the next full year.

Control of the epidemic without overwhelming the healthcare system requires coupling social distancing measures with aggressive testing to promptly identify infectious individuals and isolate them. Response capacity is critical to lift the lockdown, so that the timeline of these interventions should be carefully planned based on achieved preparedness. We consider different levels of testing capacity starting from the month of May or June. If case isolation is performed on average 1.5 day after infection and is efficient (90% reduction of contacts), we find that identifying at least 50% of all new cases would be required to rapidly reduce the burden on the healthcare system while exiting lockdown. Lower tracing capacity starting May would need to be coupled with more vigorous social distancing to keep the epidemic under control. To release constraints on the population while building capacity, a longer lockdown till June would aid releasing the pressure on the healthcare system. Also, it would be ideal to perform contact tracing and testing while the epidemic is at low activity levels. The benefit of these measures would go beyond the epidemic mitigation and extend to revising and optimizing protocols to improve case-finding and isolation – compared to the first phase of the epidemic – under more controlled conditions (reduced mixing of the population).

Fast, efficient, and large-scale contact tracing^31^ is one essential component allowing the partial release of social distancing constraints in the upcoming months. This would require digital technologies that are currently being investigated in Europe^49^ following the examples of COVID-19 response of countries in Asia^50^. Logistical constraints need to be envisioned, including large-scale and rapid diagnostic capacity, large-scale adoption of the contact tracing technology by the population^51^, uptake of recommendations, and coordination across countries to allow contact tracing across borders^49^.

The set of mild or moderate interventions considered here still impose limitations. We tested strategies allowing a larger proportion of the population to go back to work, also to partially release the huge economic pressure that lockdown generated. Global economic uncertainty is at a record high^52^, due to the fear of COVID-19 pandemic spread, income losses, and globally stalled economies because of exceptional interventions freezing production. As a side effect, lockdown has likely created a forced opportunity to re-organize certain professional activities to make telework possible and efficient at larger scale than previously foreseen. Prior to lockdown, a small fraction of Europeans practiced telework^37^. If this change of paradigm is maintained beyond emergency response, it would be extremely valuable in the medium- to long-term to aid the control of the epidemic below healthcare system saturation. Rotation of individuals working from home (e.g. every week, or every 2 weeks) can be envisioned to maintain the required social distancing levels in the community while ensuring real-life connections.

Here we consider unchanged intervention measures regarding children and seniors across all scenarios. Schools are assumed to remain closed, though reopening of certain school levels is possible under different protocols of attendance^44^. Seniors are considered to maintain a reduction of contacts through hygienic measures and physical distance, as they are especially vulnerable against COVID-19. Planning for the upcoming months under these conditions should include logistics to facilitate daily routines of the elderly beyond this phase of emergency, e.g. improving delivery of grocery and medicines, facilitating remote access to healthcare, providing learning programs for the use of technologies to stay connected, and other initiatives. Reopening of the schools in the fall/winter should be explored in the following months once the impact of these interventions will be further assessed^44^.

ICU capacity underwent a large increase in the last weeks to face the rapid surge of patients in critical conditions^16^. Capacities have been stretched in the most affected regions, and patients have also been transferred to other regions for adequate care. Exiting the current emergency, we envision that ICU capacity will be restored to lower levels for the upcoming months. If a second emergency were to occur, the system would need to be strengthened again to higher limits.

Our findings are based on the mechanistically reconstructed changes in the contact matrices that aim to reproduce the implemented social distancing measures, as done in previous works^7,12^. While reconstructing changes in the contact matrices remains arbitrary, available elements support our estimates. First, not being fitted to the lockdown period, our contact matrices reconstructed from social contact data lead to model projections in line with observations across several indicators. The fit to the epidemic trajectory, retrospectively performed once data became available, shows that a correction of only 5% in the transmissibility per contact is needed to better describe the epidemic compared to our mechanistically reconstructed matrices. This indicates that the assumptions behind the reconstruction of the matrix, affecting each age bracket in a different way, are able to capture the dynamics of the epidemic during lockdown. Such finding can improve the parameterization of similar models based on contact matrices for the study of COVID-19 epidemic in other regions or countries. At the same time, it also suggests that physical contacts were successfully avoided during lockdown, in compliance with the recommendations of health authorities. Second, our predicted reduction of 81% of the average number of contacts during lockdown is lower than the one measured in China in the cities of Wuhan (86%) and Shanghai (89%)^46^. Stricter measures were implemented in China during lockdown compared to Europe, including for example complete suspension of public transport, ban of cars from roads except for the essential services, barring residents from leaving the apartment in certain areas or limiting it to one household member few days per week, performing health checks door to door to identify and isolate ill individuals. These measures are expected therefore to have a more substantial effect on reducing the number of contacts per individual compared to social distancing measures implemented in many European countries. Indeed, under conditions measured in China, the ICU system is predicted by our model to receive less patients and clear them more rapidly than what we currently see in the data. Third, our prediction for contact reduction is close to, but larger than the empirically estimated 73% reduction of a recent social contact survey conducted in the UK during the lockdown phase^45^. Implementation of social distancing however differs in the two countries. For example, in the UK parks remain open, no self-declaration to circulate is needed, and displacements are not restricted on distance. Assuming the conditions measured in the UK, the model predicts a first peak exceeding ICU capacity. Collecting contact data in France during lockdown, as done in the UK, would allow a better measurement of the mixing patterns altered by social distancing to be compared with our synthetically reconstructed lockdown matrix. Possible changes in population adherence over time, and consequential strengthening of measures by authorities, need however to be taken into account.

Our study is affected by limitations. We did not include explicitly the effect of using masks. Evidence on seasonal coronaviruses indicate that surgical masks may reduce onward transmission^53^. Masks are largely adopted or enforced in Asia, while they just became a recommended or compulsory protection in certain areas in Europe and United States, mainly as a precautionary measure^54^. If effective, their widespread use may help decrease the risk of transmission in the community. As more epidemiological evidence accrues, this effect can be taken into account and help further alleviate control measures. We did not consider seasonal behavior in viral transmission^11,13^, because of current lack of evidence. In our simulated epidemics, multiple peaks are observed because of the implementation of social distancing interventions able to reduce the reproductive number below 1. If seasonal forcing is to be expected, the interplay with seasonality should be carefully considered in the planning of the short- and long-term control strategies^11^. Exit strategies are based on matrices including both physical and non-physical contacts. We saw that excluding physical contacts substantially contributed to the reduction of the reproductive number during lockdown. At this stage, however, large uncertainty exists on recommendations and protocols imposed by authorities to phase out lockdown. Moreover, our scenarios plan out for several months, and recommendations as well as population adoption may strongly change over this long time period. Findings reported in the main text therefore corresponds to a conservative choice. If physical contacts are avoided for several months, the epidemic would go locally extinct. Treatment of COVID-19 patients improved over time, as documented by lower probabilities for requiring intensive care and shorter durations in ICU (Additional file 1). These data became available at the time of revision and were not included in the analysis. The more efficient management of COVID-19 patients is expected to reduce the burden in the upcoming months.

We tested two values for the probability of being asymptomatic, as there still exist large uncertainty^20– 23^. Few studies investigated the clinical progression of symptoms over time until viral clearance. Additional household studies now launched in Europe will help providing a better understanding of the presence of asymptomatic cases and their contribution to transmission. Evidence so far seems to indicate that this fraction may be low^23^, therefore we presented in the main paper results for *p*_*a*_ =0.2. Estimates of age-specific severity and case fatality rates are still rapidly evolving and often vary across countries due to different surveillance systems and testing protocols in place. Here we used estimates of age-specific severity informed from a model-based analysis on individual-level data from China and other countries^19^. Rates describing the hospitalization duration and outcome of a patient were based on French data^28^. Infection fatality ratios estimated by our model are consistent with estimates provided in Ref.^19^. We do not consider here data on comorbidities that will alter hospitalization and fatality rates. Large-scale testing in France will allow us to robustly estimate age-specific hospitalization rates to better inform the model.

Concurrently to our work, other studies became available that assessed the epidemic in France^35,48^. All studies independently produced similar estimates characterizing the epidemic and the effectiveness of lockdown, providing a consensus of evaluations despite modeling discrepancies (e.g. equal transmissibility across asymptomatic and symptomatic infectious individuals in Ref.^35^, or 90% reduction in transmissibility of asymptomatic cases in Ref.^48^, compared to our 45% reduction informed by prior modeling work^8^). However, each focused on different aspects, specifically to evaluate the impact of lockdown and estimate population immunity^35^, or to estimate the total number of averted ICU admissions and deaths due to lockdown^48^, whereas no study proposed exit strategies and their evaluation while on lockdown.

Our results are based on data from Île-de-France, currently one of the most affected region by the COVID-19 epidemic in the country, and not directly applicable to other regions. Few differences in the results due to variations of age profile across regions are to be expected^55^. The most relevant changes will however result from the different epidemic phase experienced by each region at the moment of nationwide application of the lockdown. Overall, findings on exit strategies remain valid, but more specific interventions, differentially targeting the regions, may be envisioned.

## CONCLUSIONS

France, as many other countries in the world, implemented a nationwide lockdown to curb the dramatic increase in the number of patients in critical conditions. Assessing the impact of lockdown and identifying the optimal strategies to manage the health crisis beyond lockdown is of critical importance. Substantial social distancing will be needed in the upcoming months due to the currently low population immunity. Given the features of COVID-19 pandemic, extensive case-finding and isolation following lockdown are required to progressively lower the intensity of current interventions and avoid that the healthcare system exceeds saturation. Response planning needs to urgently prioritize the logistics and capacity for these interventions.

## Data Availability

All data are available at the references cited.

## SUPPLEMENTARY INFORMATION

### Additional file 1

Supplementary Material composed of Supplementary results, Tables S1 and S2, and Figures S1-S10.

## Acknowledgments

We thank Chiara Poletto, Fabrice Carrat, Cecile Souty, Niel Hens, Pietro Coletti, Lander Willem, Nicola Scarpa, Guillaume Beraud, Juliette Paireau and Sante publique France for useful discussions.

## Authors’ contributions

VC conceived and designed the study. LDD, CES, PYB collected and analysed the data. LDD, GP performed the simulations and numerical analysis. LDD, GP, CES, PYB, VC interpreted the results. VC drafted the Article. All authors contributed to the writing of the final version of the Article. All authors read and approved the final manuscript.

## Availability of data and materials

All data is available at cited sources.

## Ethics approval and consent to participate

Not applicable.

## Consent for publication

The funders had no role in study design, data collection, data analysis, data interpretation, writing of the manuscript, and decision to submit. The corresponding author had full access to all the data in the study and had final responsibility for the decision to submit for publication.

## ADDITIONAL FILE 1

### 1. MODEL PARAMETERS

#### 1.1. Compartmental model

**Table S1.**
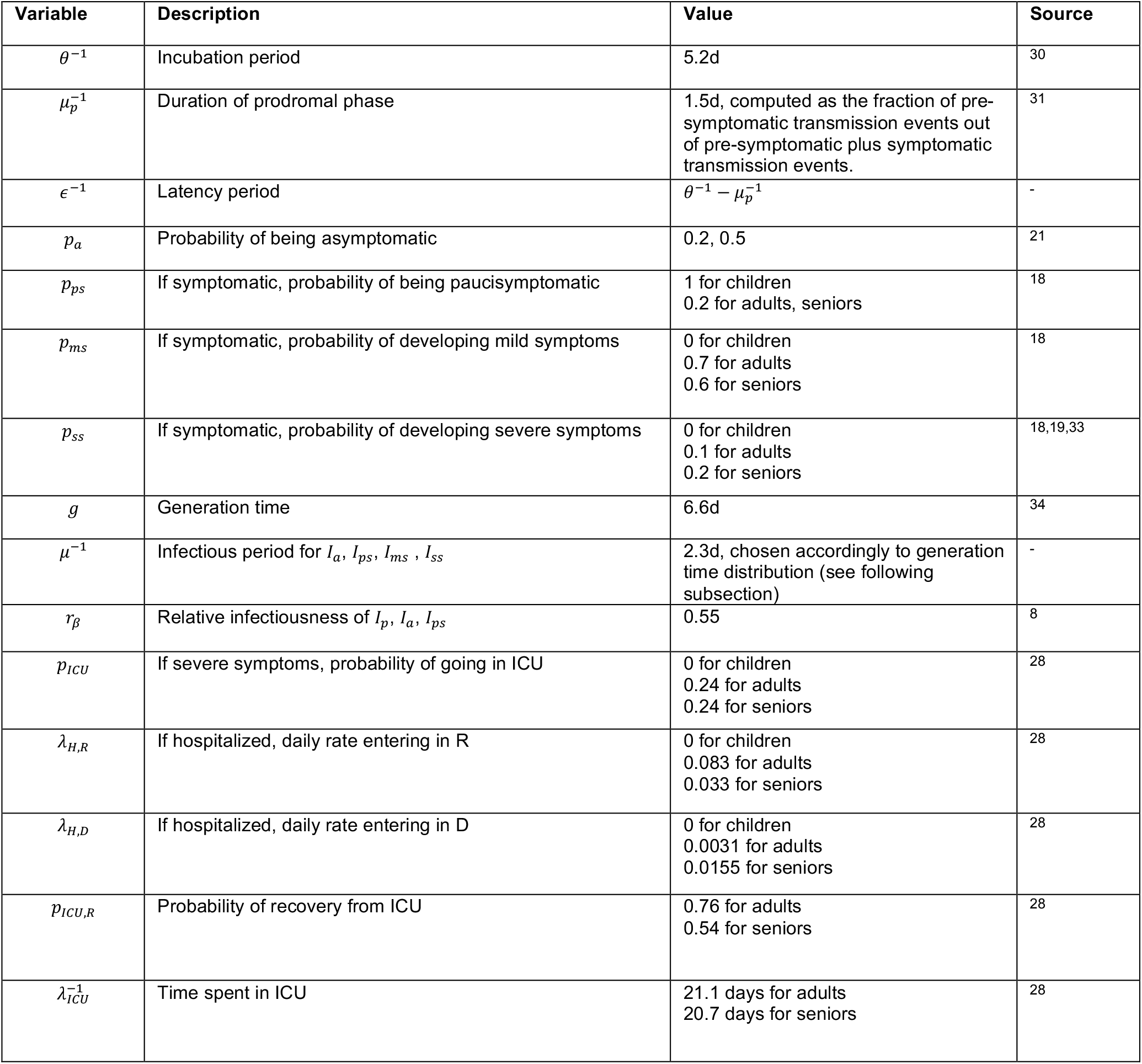
Parameters, values, and sources used to define the compartmental model

| Variable | Description | Value | Source |
| --- | --- | --- | --- |
| $\theta^{-1}$ | Incubation period | 5.2d | 30 |
| $\mu_p^{-1}$ | Duration of prodromal phase | 1.5d, computed as the fraction of pre-symptomatic transmission events out of pre-symptomatic plus symptomatic transmission events. | 31 |
| $\epsilon^{-1}$ | Latency period | $\theta^{-1} - \mu_p^{-1}$ | - |
| $p_a$ | Probability of being asymptomatic | 0.2, 0.5 | 21 |
| $p_{ps}$ | If symptomatic, probability of being paucisymptomatic | 1 for children<br>0.2 for adults, seniors | 18 |
| $p_{ms}$ | If symptomatic, probability of developing mild symptoms | 0 for children<br>0.7 for adults<br>0.6 for seniors | 18 |
| $p_{ss}$ | If symptomatic, probability of developing severe symptoms | 0 for children<br>0.1 for adults<br>0.2 for seniors | 18,19,33 |
| $g$ | Generation time | 6.6d | 34 |
| $\mu^{-1}$ | Infectious period for $I_a$, $I_{ps}$, $I_{ms}$, $I_{ss}$ | 2.3d, chosen accordingly to generation time distribution (see following subsection) | - |
| $r_\beta$ | Relative infectiousness of $I_p$, $I_a$, $I_{ps}$ | 0.55 | 8 |
| $p_{ICU}$ | If severe symptoms, probability of going in ICU | 0 for children<br>0.24 for adults<br>0.24 for seniors | 28 |
| $\lambda_{H,R}$ | If hospitalized, daily rate entering in R | 0 for children<br>0.083 for adults<br>0.033 for seniors | 28 |
| $\lambda_{H,D}$ | If hospitalized, daily rate entering in D | 0 for children<br>0.0031 for adults<br>0.0155 for seniors | 28 |
| $p_{ICU,R}$ | Probability of recovery from ICU | 0.76 for adults<br>0.54 for seniors | 28 |
| $\lambda_{ICU}^{-1}$ | Time spent in ICU | 21.1 days for adults<br>20.7 days for seniors | 28 |

#### 1.2. Generation time distribution

The generation time distribution in a compartmental epidemic model can be computed thanks to the theory developed by Svensson^32^. Let *X* and *Y* be the random variables describing the latency period and the infectious period, respectively. Then the distribution of the generation time is the result of the convolution *g *h*_*s*_, with *g* being the probability density function of *X* and

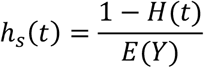

where *H* is the cumulative distribution function of *Y*, and *E*(*Y*) is the mean.

In the compartmental model under consideration (Figure 2), we have that *X*is exponentially distributed with rate *ε*, and *Y* is the sum of two exponentially distributed random variables (prodromic phase and infectious period, with rate *μ*_*p*_ and *μ* respectively). Computations show that the corresponding generation time distribution is

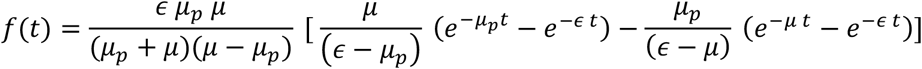

Given the values of *∈* and *μ*_*p*_ informed from the literature (Table S1), we choose *μ* so that the mean of the generation time equals to 6.6 days. The shape of the distribution is displayed in Figure S1 and it closely resembles a gamma distribution with mean 6.6 and shape parameter 1.87, estimated in Ref.^34^

**Figure S1.**
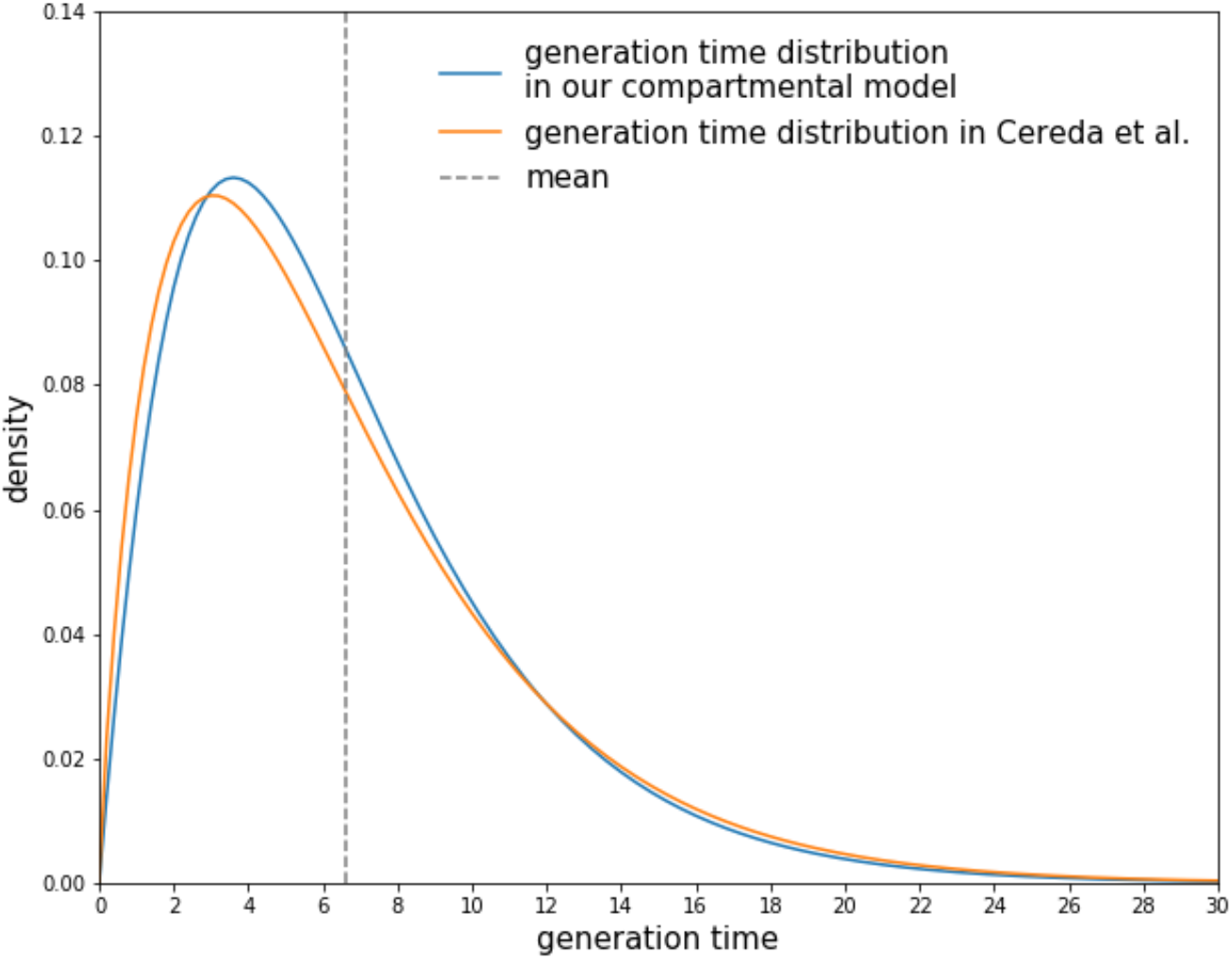
Distribution of the generation time. The generation time distribution corresponding to our compartmental model (blue) in comparison with the distribution estimated in Ref.^34^ (orange).

#### 1.3. Estimation of within-hospital parameters

We fit data on patient trajectories recorded in Île-de-France hospitals after admission up to April 5, 2020. Data consisted of age, sex, date of hospital admission and subsequent dates of discharge or death, and, when relevant, dates of entering/leaving the ICU. We fit mixture and competing risks models to time to event data, taking into account censoring due to patients being still in the hospital at the time of analysis. We used exponential distributions for time to event data to match the hypotheses of the compartmental epidemic model.

First, we model time from admission to entering the ICU or being discharged/dead for those who do not go to the ICU. Write T for the time to the first of the 3 following events: entering the ICU, being discharged alive or dying in the hospital. T is modelled as a mixture of 2 exponential distributions: T ∼ *π*_*ICU*_ *Exp*(*λ*_*ICU*_)+(1 − *π*_*ICU*_) *Exp* (*λ*_*H*_), where *π*_*ICU*_ is the probability to go to the ICU, and *λ*_*ICU*_,*λ*_*H*_ are the rates of the exponential distributions. The second exponential describes time spent in the hospital by those who don’t go the ICU subject to competition of 2 outcomes, discharge or death. Therefore, *λ*_*H*_ = *λ*_*DIS*_ + *λ*_*DTH*_ where *λ*_*DIS*_ is the rate of discharge and *λ*_*DTH*_ the rate of death. The average time spent in the hospital is 1/*λ*_*H*_, and the probability of being discharged alive is *λ*_*DIS*_/(*λ*_*DIS*_ + *λ*_*DTH*_). Therefore, the likelihood of a patient trajectory observed up to time t with final status s (comprising still hospitalized - HOS, admitted to ICU - ICU, discharged alive - DIS, dead - DTH) is given by:

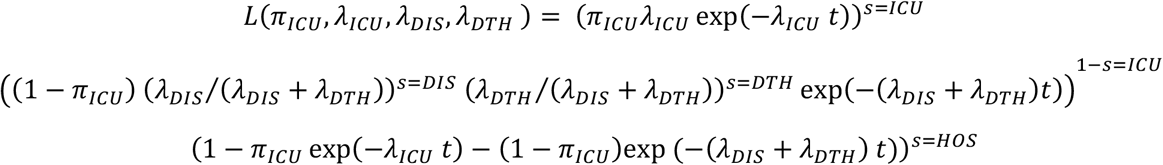

The first line is for patients going to the ICU, the second line for those being discharged alive or dead and the third line for patients who were censored because they were still in the hospital.

Likewise, we fit time to discharge or death after admission to the ICU using a competing risk approach with exponential parameters *µ* for being discharged alive or dead; the likelihood is therefore:

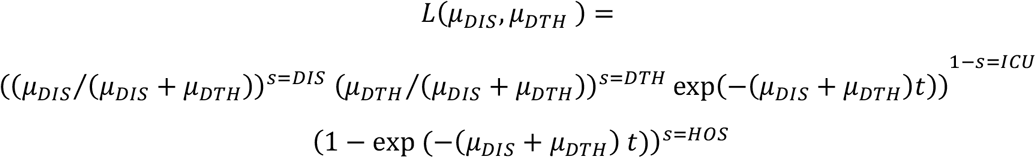

As the data is rounded to the nearest day, we discretized the exponential distributions in the likelihood. All models were fitted at maximum likelihood using the software R.

Estimates up to April 5, 2020 were also compared to values estimated in the period April 5-26 to assess possible changes in the management of COVID-19 patients at the hospital. We report the values in Table S2. These estimates are discussed in the main paper, but are not included in the analysis, as they became available at a later time.

**Table S2.** Estimates of within-hospital parameters in two different periods of time.

|  | March 1 – April 5, 2020 | April 5 – 26, 2020 |
| --- | --- | --- |
| $p_{ICU}$ | 0.24 for adults, seniors | 0.16 for adults, seniors |
| $\lambda_{H,R}$ | 0.0832 for adults<br>0.0328 for seniors | 0.0834 for adults<br>0.0330 for seniors |
| $\lambda_{H,D}$ | 0.0031 for adults<br>0.0155 for seniors | 0.0023 for adults<br>0.0115 for seniors |
| $p_{ICU,R}$ | 0.76 for adults<br>0.54 for seniors | 0.84 for adults<br>0.64 for seniors |
| $\lambda_{ICU}^{-1}$ | 21.1 days for adults<br>20.7 days for seniors | 15.0 days for adults<br>16.6 days for seniors |

#### 1.4. Mixing matrices

Here we report the matrices computed for all interventions tested, compared to the baseline scenario.

**Figure S2.**
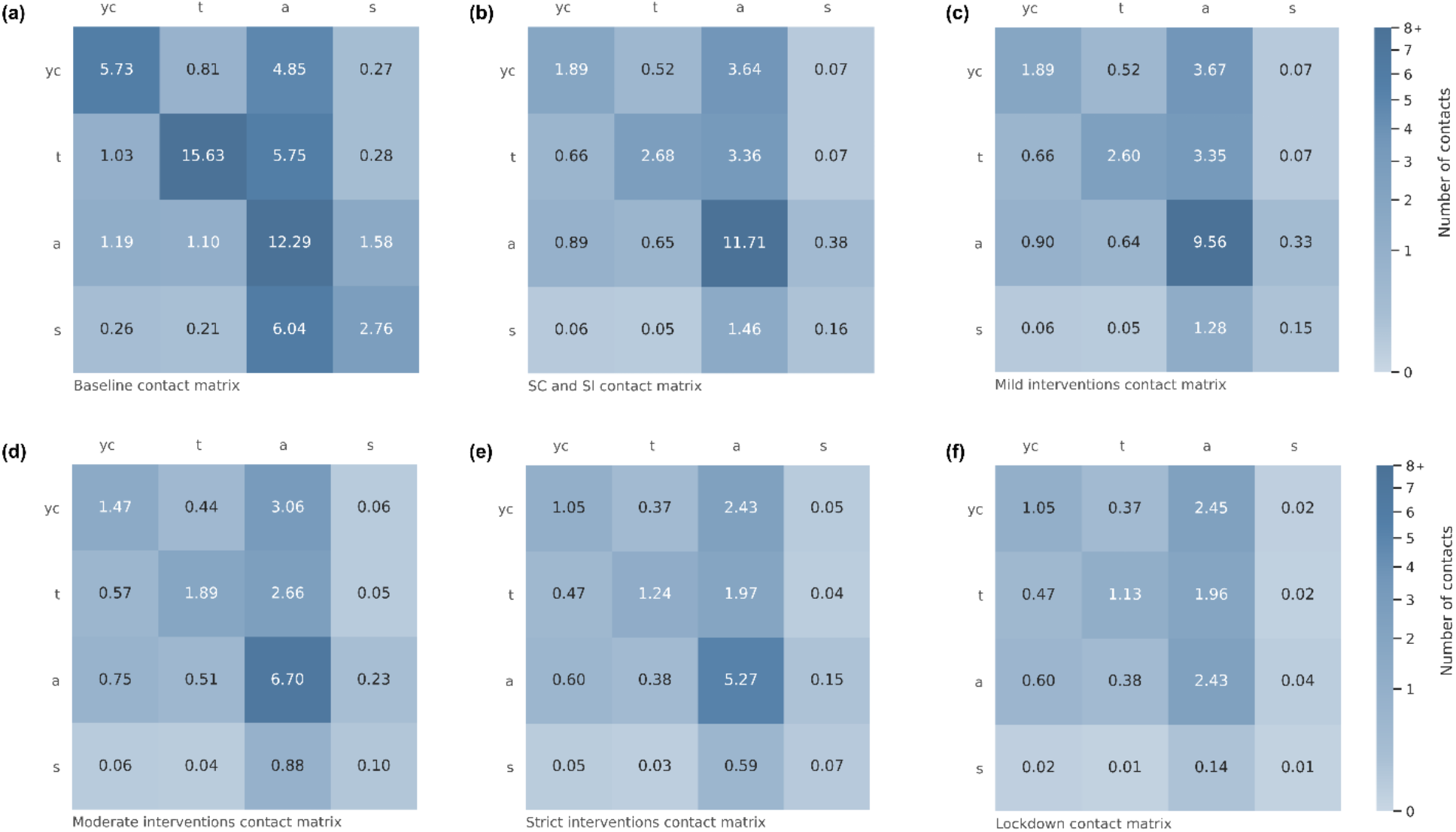
Mixing matrices for the baseline and all social distancing interventions tested. (a) Baseline contact matrix (b) School closure and senior isolation contact matrix (c) Mild interventions contact matrix (d) Moderate interventions contact matrix (e) Strict interventions contact matrix (f) Lockdown contact matrix.

### 2. MODEL CALIBRATION

The model was calibrated to hospital admission data through a maximum likelihood approach. The likelihood function is of the form

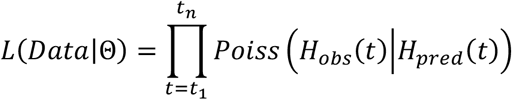

where Θindicates the set of parameters to be estimated, *H*_*obs*_(*t*) is the observed number of hospital admissions on day *t, H*_*pred*_(*t*) is the number of hospital admissions predicted by the model on day *t, Poiss* (· |*H*_*pred*_(*t*)) is the probability mass function of a Poisson distribution with mean *H*_*pred*_(*t*), and [*t*_1_,*t*_*n*_] is the time window considered for the fit. We fit the transmission rate per contact before lockdown and the starting date of the simulation, considering a time window ranging from March 1 to March 23, 2020. Hospital admissions in the interval March 17-23 were included as still not affected by lockdown, due to delay between date of infection and date of hospitalization (∼1 week). The resulting estimate of the transmission rate is 0.0791, with 95% CI [0.0769, 0.0806], corresponding to *R*_*a*_=3.18 [3.09, 3.24].

Our model predictions were also compared to the ones obtained by fitting the model to hospital admission data during lockdown. We chose the period April 13-May 10 to avoid the initial fluctuations due to the implementation of lockdown. By doing that, we obtain an estimated transmission rate at 0.0833, corresponding to a 5.3% increase with respect to the transmission rate prior to lockdown.

The number of hospital admissions, ICU admissions and ICU occupancy over time for Ile-de-France (Figure 4a, Figure 5) are available in an open access repository (https://docs.google.com/spreadsheets/d/17Q5BlJw2N6b5uf8T2E3leTul91tAyqOvKhymvPDyhHk/edit?usp=sharing). They are part of the SIVIC database maintained by the Agence du Numérique en Santé and Santé Publique. The same online repository also contains the incidence of clinical cases estimated from sentinel and virological surveillance by Réseau Sentinelles (Figure 4b).

### 3. ADDITIONAL RESULTS

#### 3.1. Consultation rate throughout lockdown

**Figure S3.**
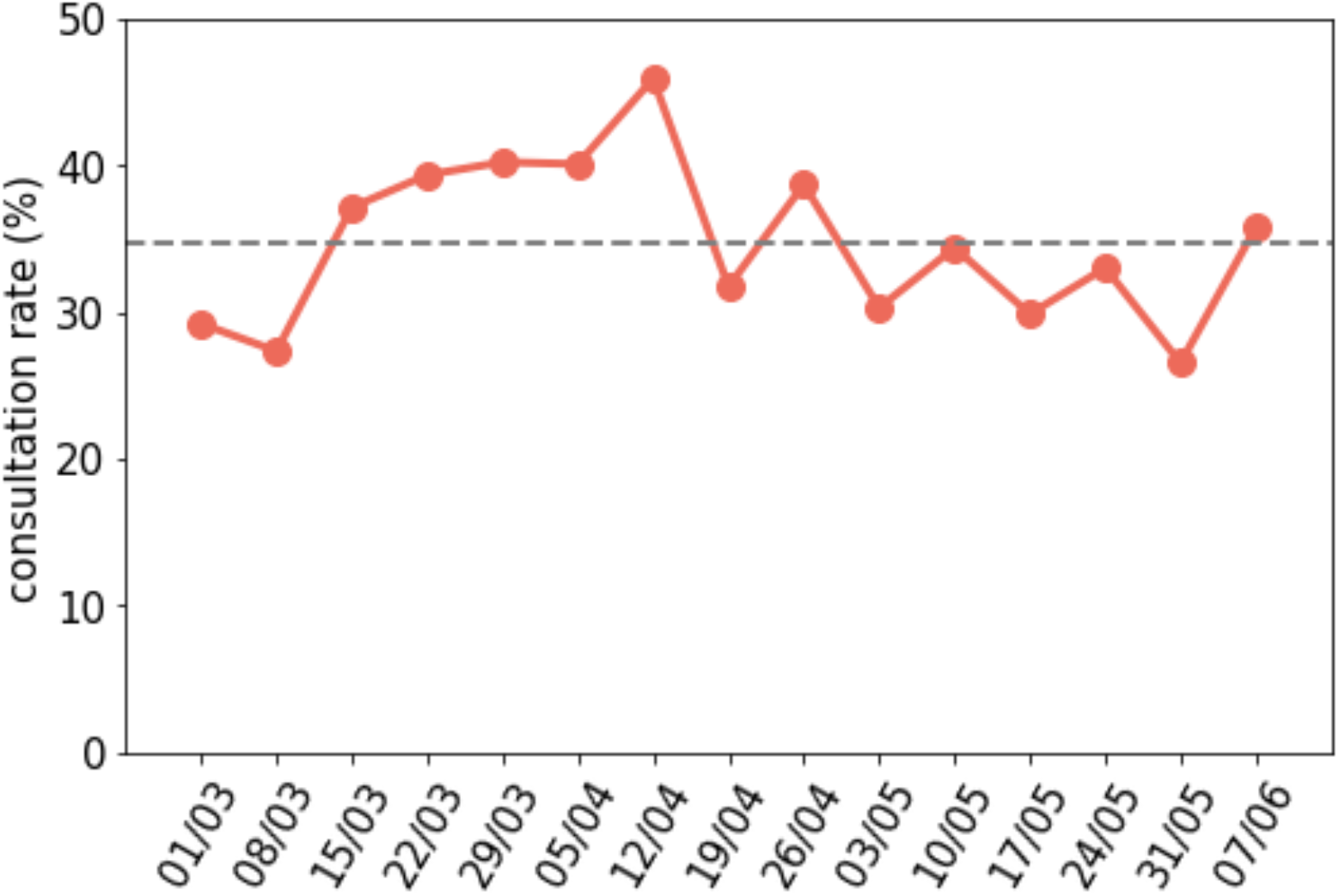
Consultation rate during lockdown estimated from crowdsourced data^38^.

#### 3.2. Lockdown lifted at the beginning of May

**Figure S4.**
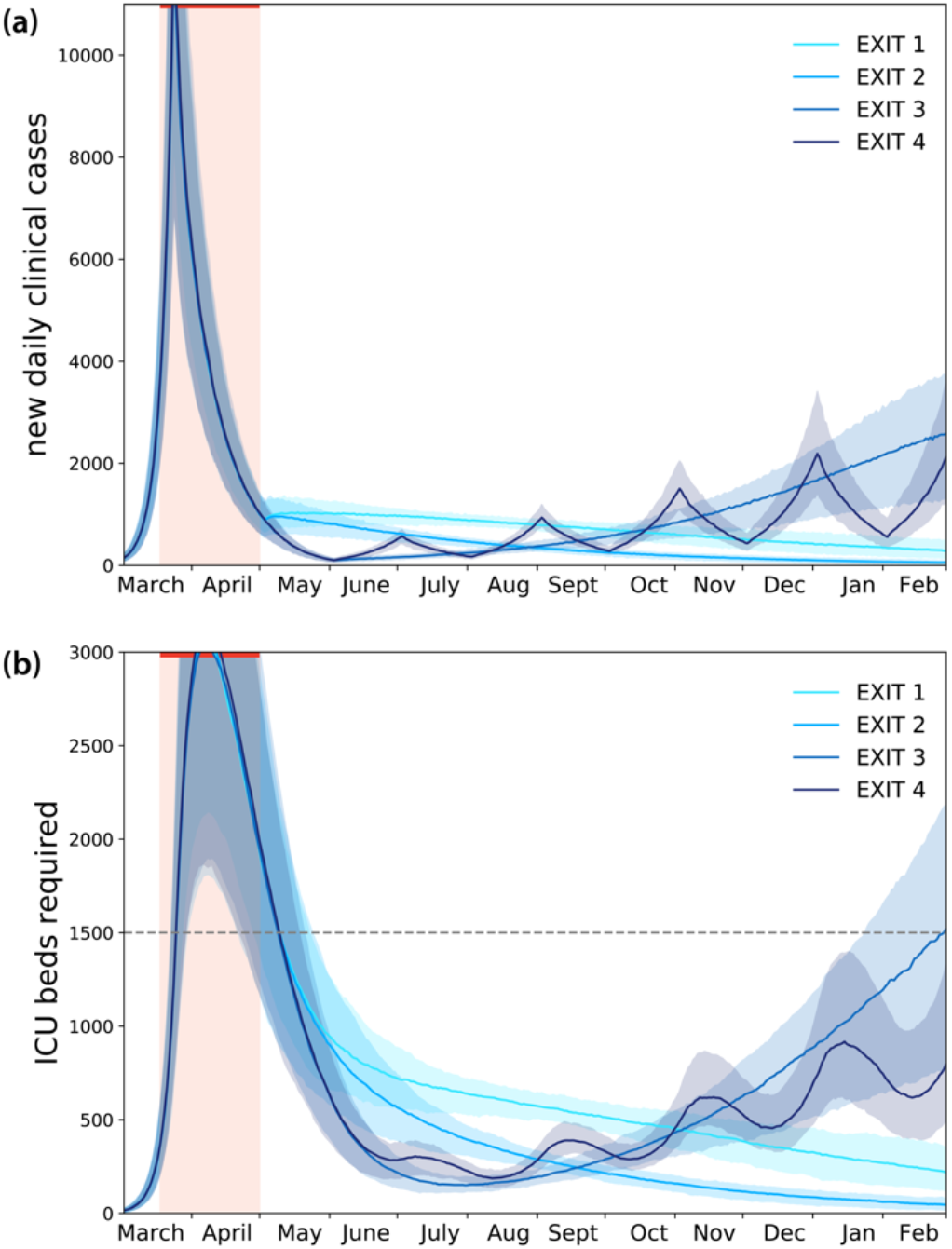
Simulated impact of lockdown and exit strategies with large-scale testing and case isolation, once lockdown is lifted on May 1. (a) Simulated daily new number of clinical cases assuming the progressive exit strategies illustrated in Figure 3. (b) Corresponding demand of ICU beds.

#### 3.3. Physical contacts avoided throughout the exit strategies

**Figure S5.**
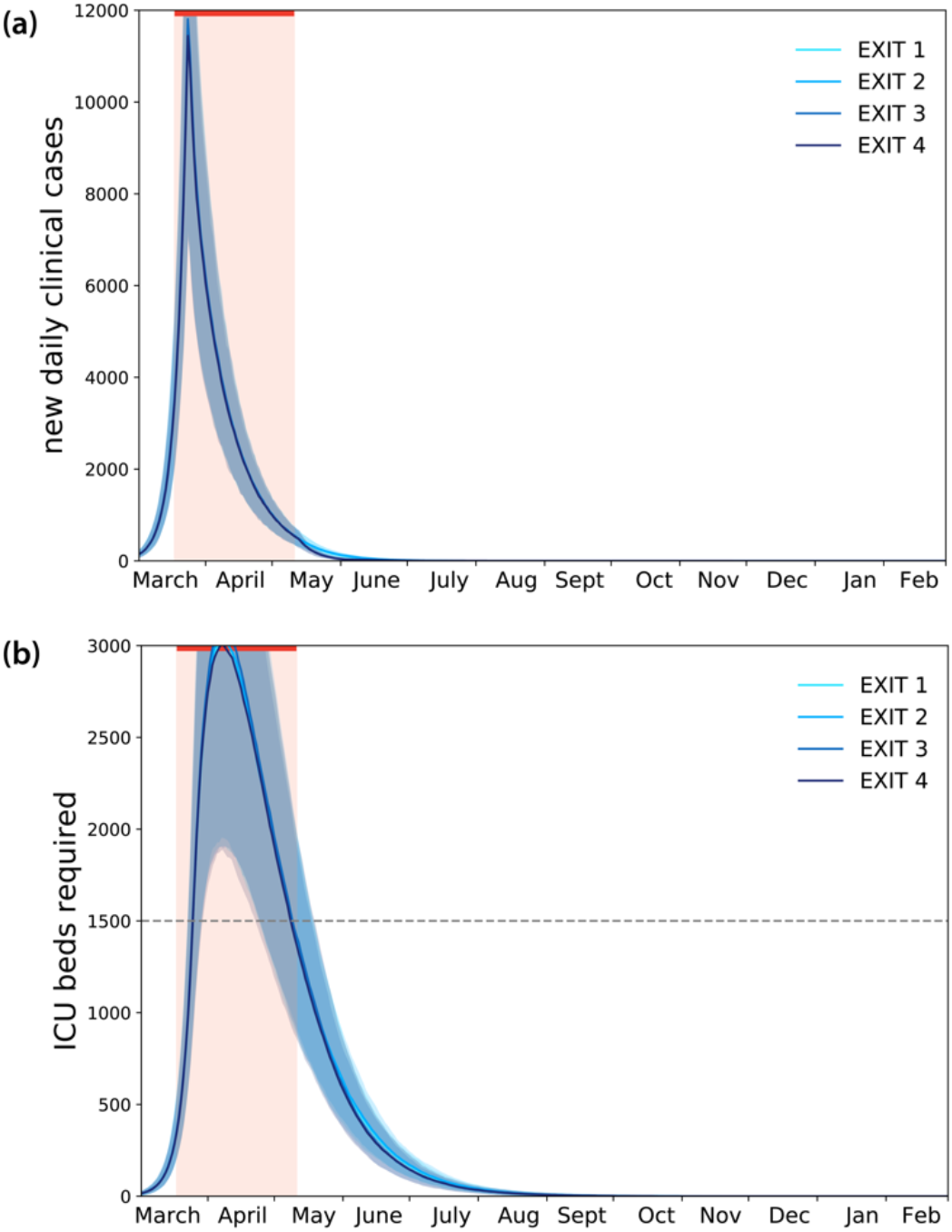
Simulated impact of lockdown and exit strategies with large-scale testing and case isolation, if physical contacts are avoided throughout the exit strategies. (a) Simulated daily new number of clinical cases assuming the progressive exit strategies illustrated in Figure 3. (b) Corresponding demand of ICU beds.

### 4. SENSITIVITY ANALYSIS

#### 4.1. Probability of being asymptomatic 50%

Here we report the numerical results obtained assuming a higher probability of being asymptomatic (*p*_*a*_ =0.5) compared to the main paper (*p*_*a*_ =0.2) (Figures S6-S8).

**Figure S6.**
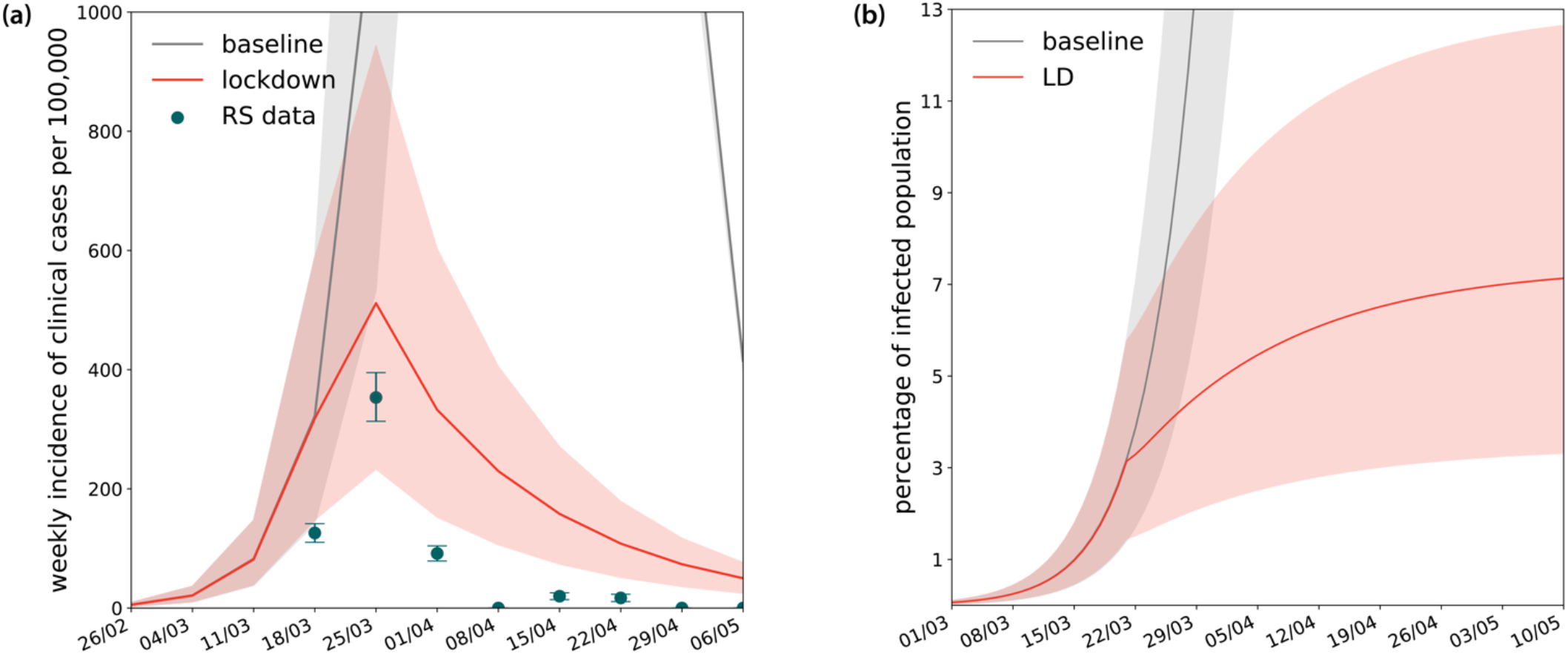
Estimates of weekly incidence and percentage of population infected. (a) Simulated weekly incidence of clinical cases (mild and severe) compared to estimates of COVID-19 positive cases in the region provided by syndromic and virological surveillance (Reseau Sentinelles (RS) data)^42^. (b) Simulated percentage of population infected over time. Results are shown for *p*_*a*_=0.5. Shaded areas correspond to 95% probability ranges.

**Figure S7.**
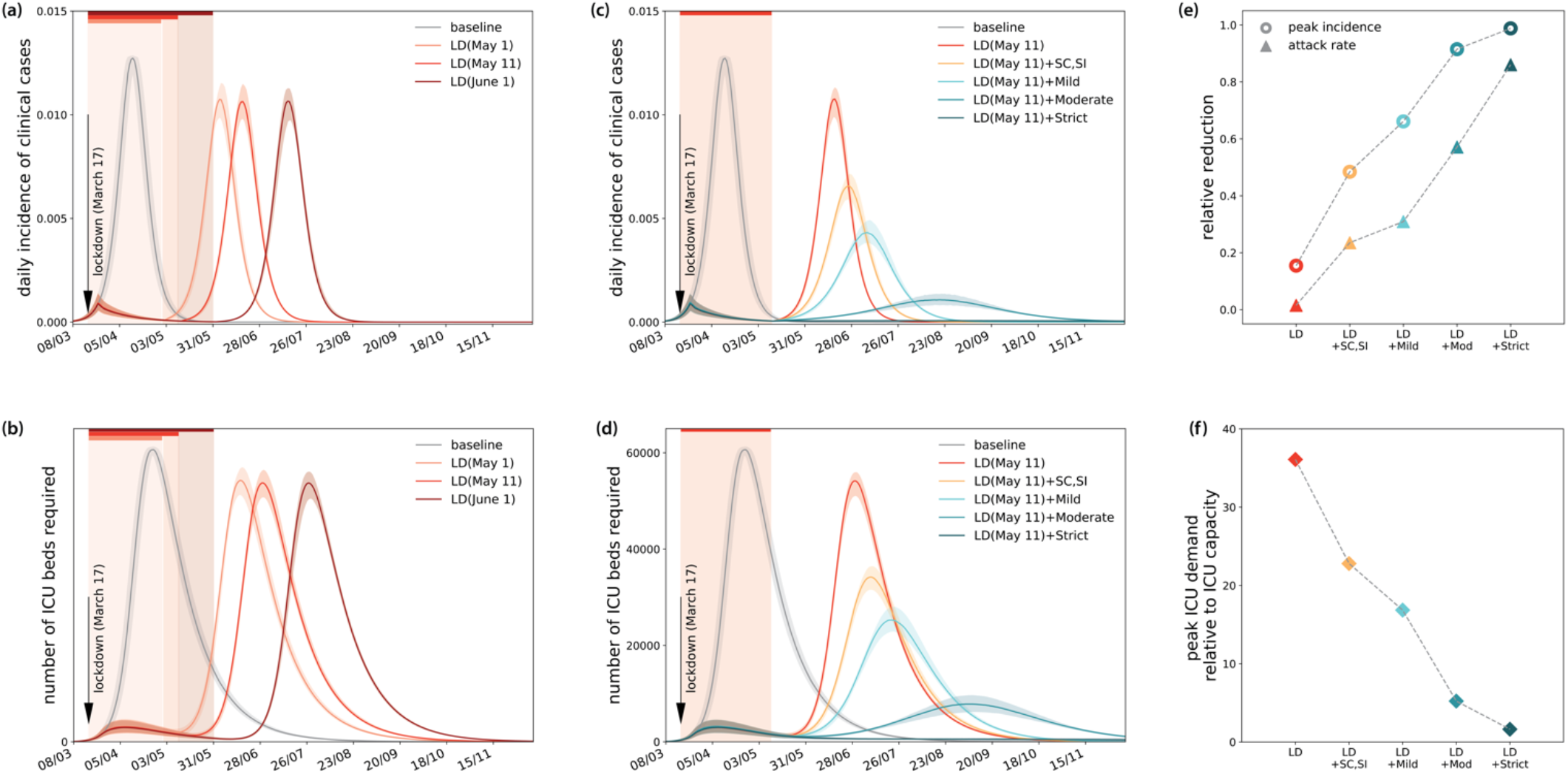
Simulated impact of lockdown of different durations and exit strategies. (a) Simulated daily incidence of clinical cases assuming lockdown till end of April, May 11, end of May. (b) Corresponding demand of ICU beds. (c) Simulated daily incidence of clinical cases assuming lockdown till May 11, followed by interventions of varying degree of intensity. (d) Corresponding demand of ICU beds. (e) Relative reduction of peak incidence and epidemic size after 1 year for each scenario. (f) Peak ICU demand relative to restored ICU capacity of the region (1,500 beds). In all panels, the color code is as in Table 1, and scenarios are identified as reported in Figure 3 in the main paper. Baseline scenario corresponds to no intervention. Results are shown for *p*_*a*_=0.5.

**Figure S8.**
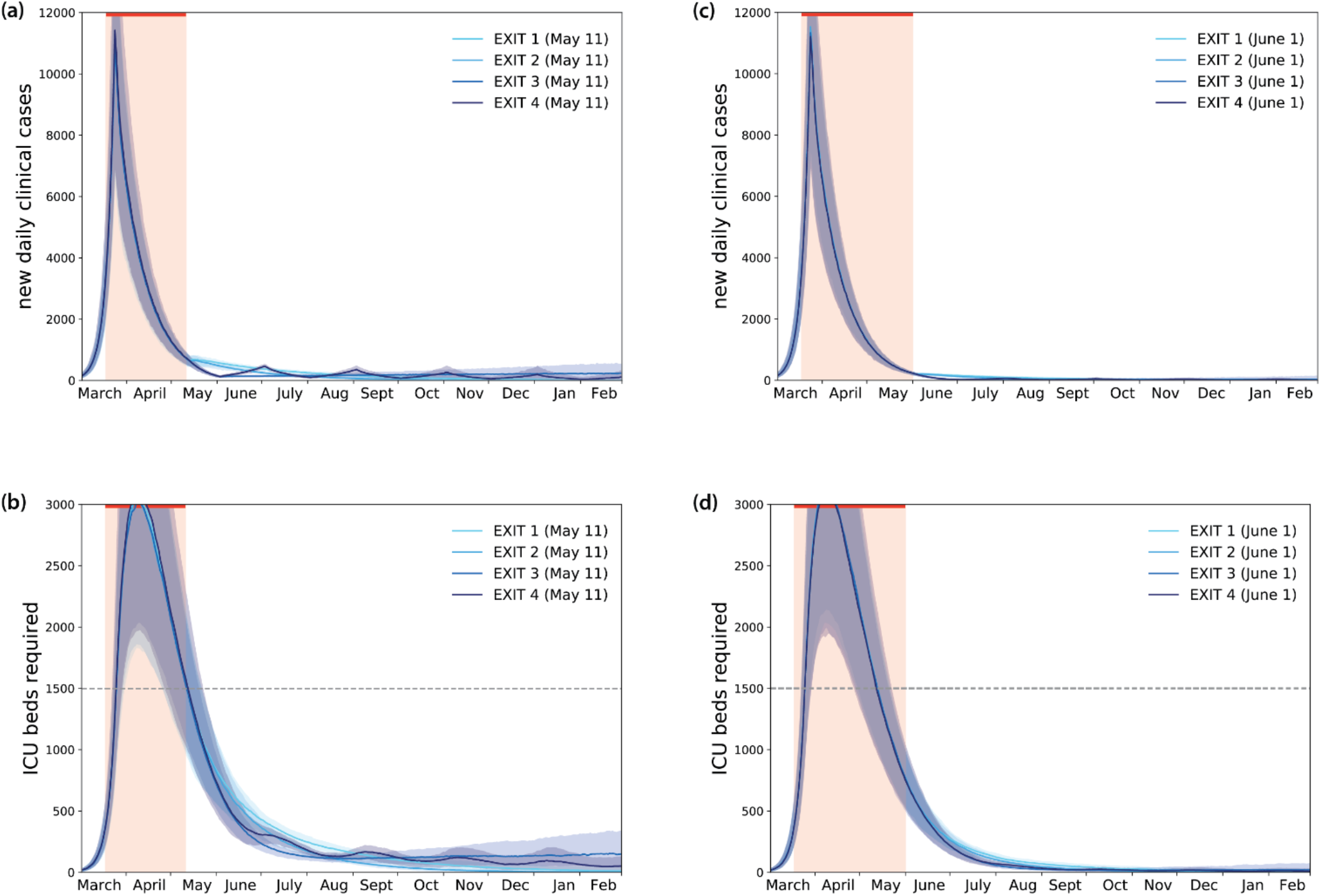
Simulated impact of lockdown and exit strategies with large-scale testing and case isolation. (a) Simulated daily new number of clinical cases assuming the progressive exit strategies illustrated in Figure 3. (b) Corresponding demand of ICU beds. (c) as in (a) with strategies implemented 1 month after, i.e. keeping a lockdown till the end of May. (d) Corresponding demand of ICU beds. Results are shown for *p*_*a*_=0.5.

#### 4.2. Relative susceptibility of children

**Figure S9.**
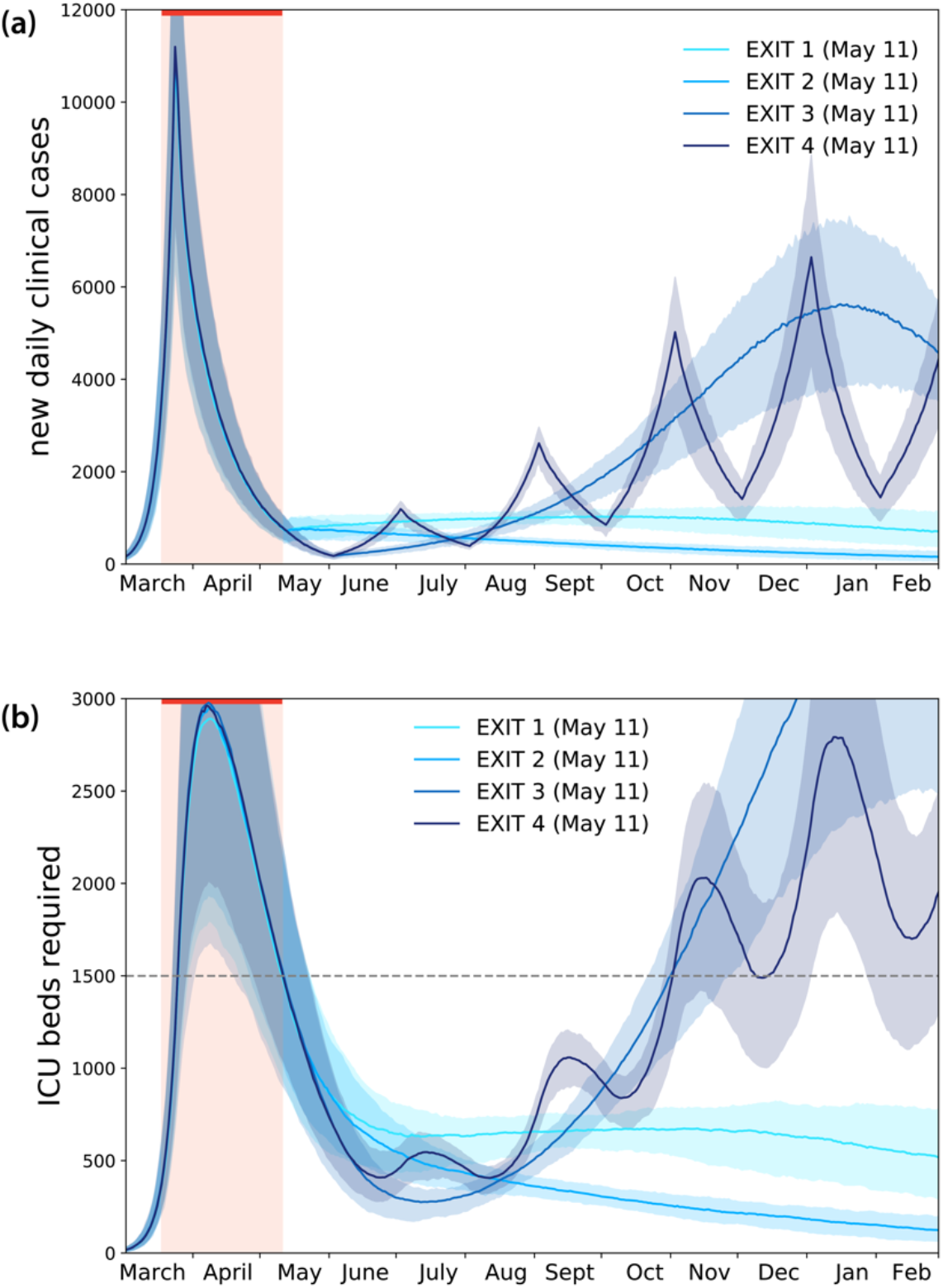
Simulated impact of lockdown and exit strategies with large-scale testing and case isolation, assuming that children (under 19 years of age) are 50% susceptible compared to adults. (a) Simulated daily new number of clinical cases assuming the progressive exit strategies illustrated in Figure 3. (b) Corresponding demand of ICU beds.

#### 4.3. Relative infectivity of younger children

**Figure S10.**
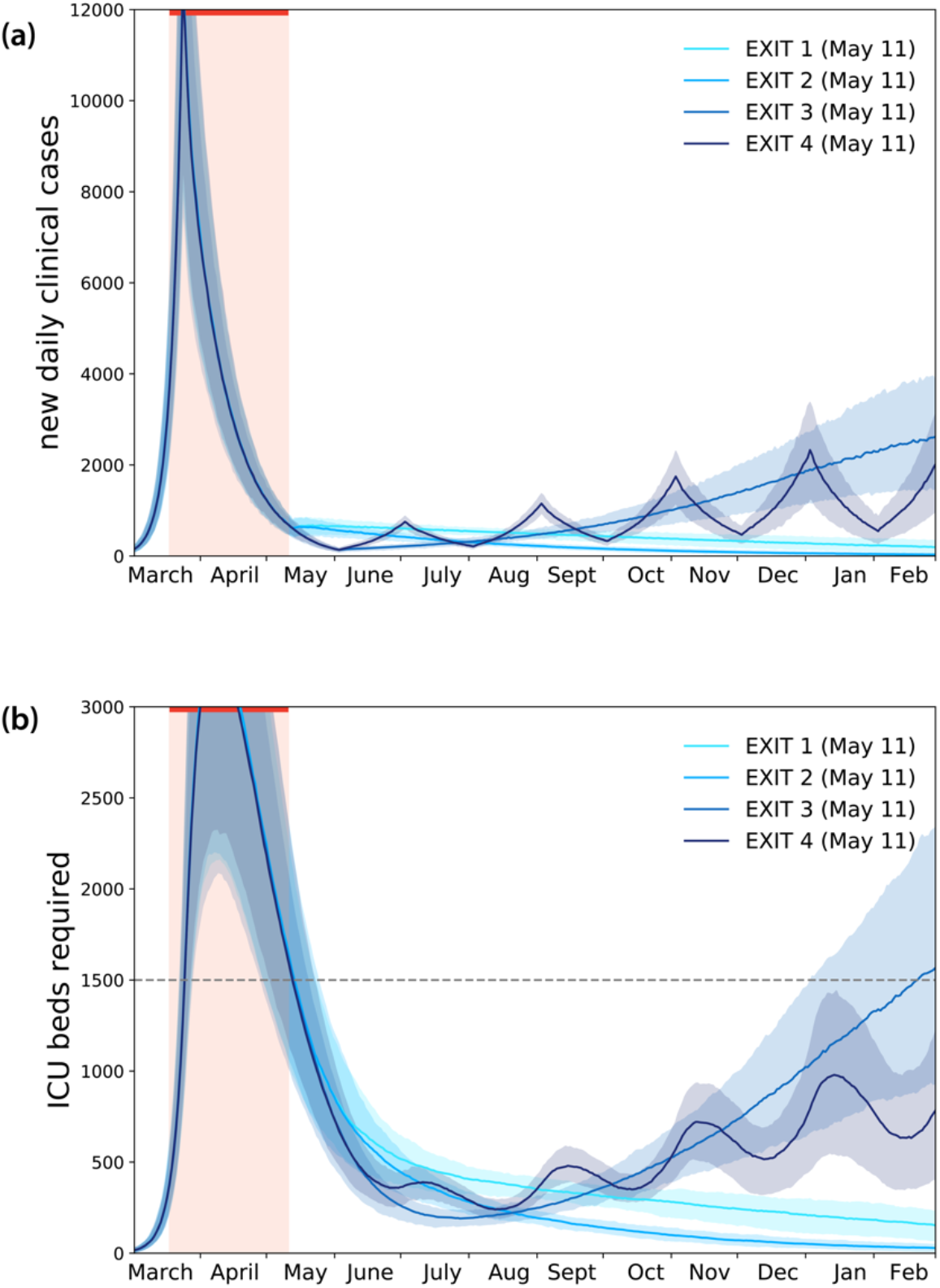
Simulated impact of lockdown and exit strategies with large-scale testing and case isolation, assuming that younger children (below 10 years of age) are 50% less infectious than adolescents^44^. (a) Simulated daily new number of clinical cases assuming the progressive exit strategies illustrated in Figure 3. (b) Corresponding demand of ICU beds.

